# In Vivo Spatial Transcriptomics for Bleeding-free Profiling Human Internal Organs

**DOI:** 10.64898/2026.07.06.26357355

**Authors:** Hailiang Sun, Feng Guo, Xinzhe Zhao, Youyang Wan, Xijian Zhang, Jiachen Sun, Xingdao He, Baowen Gai, Chuxiao Xiong, Yutao Ma, Jin Qu, Pengyu Li, Feng Gao, Xi Zhao, Xianglin Ji, Zhengbao Yang, Lung-Yi Mak, Desmond Yat Hin Yap, Jia Ke, Peng Shi

## Abstract

Despite the significant technical advancement in spatial transcriptomics, its clinical usage is largely untapped. Here, we develop an integrated system, ENDO-Genome, for minimally invasive in-body transcript sampling to facilitate live spatial transcriptomic analysis of human internal organs. This is achieved by integrating a nanoarrayed biochip with existing endoscope to perform pressure-sensor-calibrated “Touch & Go” RNA extraction directly from human internal organs, including the highly vascularized liver or kidney, without the need for tissue biopsy, voiding any bleeding risks. By a demonstration using gastrointestinal endoscopy, multiplexed landscape of 55 mRNA transcripts was obtained from multiple locations of human intestinal tract via a 5-minute operation in routine examinations. Benefiting from a sequencing-free approach, each assay costs less than 10 US dollars. For the clinical study involving 15 Crohn’s disease (CD) patients, no complication case was reported out of 47 ENDO-Genome operations, showcasing the gentle deposition and excellent safety of the technique. The live spatial transcriptomics provides direct *in vivo* pictures of the heterogenous spatial transcriptional programs underlying CD pathological response at different intestinal locations, revealing distinct ileal phenotypes. This is manifested by unique microscale scattering of inflammation gene clusters, along with the discovery of a tissue-specific cooperative mechanisms between inflammation and RNA methylation regulations at single- or multi-cell scales.

## Introduction

Spatial transcriptomics measures the gene expression within the spatial context of intact or processed tissues. The technology provides a map of gene expression at tissue-scale with a resolution approaching single-cell level, offering insights into the complex interactions among different cell populations ^1, 2^. Over the past decade, significant progresses have been made in developing spatial transcriptomic technologies with improved the resolution, sensitivity, and accessibility ^3^. Techniques like Slide-seq ^4^, STARmap ^5^, and Genomics Visium ^6^ provide the capability to analyse large tissue sections. Alternatives based on *in situ* hybridization allow researchers to achieve single-cell resolution, providing more detailed spatial maps of gene expression ^7^. More recently, more affordable solutions are emerging to further push the boundaries of spatial transcriptomics ^8, 9^. There is also a growing trend of integrating spatial transcriptomics with other omics technologies for comprehensive understanding of cellular functions and interactions ^10, 11^.

Along with the technology innovations, spatial transcriptomics has been increasingly used to understand tissue architecture from a transcriptomics perspective ^3, 12^, to study developmental biology ^13, 14^, to investigate disease mechanism based on spatial cellular heterogeneity ^15, 16^, and to help the clinical diagnostics and treatment ^17, 18^. While undergoing fast development, existing spatial transcriptomic technologies only apply to extracted tissues, which require extensive pre-processing steps involving complex spatially encoded RNA extraction. This prohibits the usage of most existing techniques directly in human body for *in vivo* profiling of internal organs to provide insights into spatially resolved genetic or epigenetic organizations.

Particularly, tissue biopsies are typically used in clinical practices to extract samples for diagnostic assays, including spatial transcriptomics ^19^. While most biopsies are routine and safe, bleeding can occur and may vary in severity depending on multiple factors, including the organ vasculature, surgical procedure and patient condition ^20^. For example, liver, kidneys, lungs, and other highly vascularized organs have much higher risks of bleeding complications ^21, 22^. The issue gets more concerned for patient on antithrombotic agents or with coagulation disorders. On the other hand, *in vivo* spatial transcriptomics to be performed in a living organism, can bypass the biopsy procedure by a direct access of native physiology environments, which not only promise a bleeding-free operation but also minimize tissue handling for reduced artefacts and enable accurate capture of dynamic transcriptomic events. This is especially important for studying rapid or transient biological events, such as epigenetics associated targets ^23^. Also, longitudinal studies can be performed to observe transcriptomics changes over time within the same organ, providing information regarding developmental processes, disease progression, and treatment responses, which shows the great clinical relevance of in-body profiling techniques as a minimally invasive diagnostic tool.

Despite the exciting frontier, technical innovation for *in vivo* spatial transcriptomics comes with significant challenges. Biocompatilbity and patient safety are the foremost concerns. Any devices must be designed to minimize invasiveness to reduce tissue damage and patient risks. Designing probes that can efficiently access and extract the target molecules, such as RNAs, from the internal tissues is not easy. achieving the resolution of similar single-cell level in a living organism as being demonstrated in the *ex vivo* systems is extremely challenging. Last but not least, cost-effectiveness and scalability are also critical for widespread clinical use.

To address these challenges, in this study, we develop an ENDO-Genome system for minimally invasive in situ spatial transcriptomic analysis of human internal organs with spatially resolved single-cell resolution. A nanoprobe structured gene chip is integrated with existing surgical tools, such gastrointestinal endoscopy system or biopsy needles, for accessing the superficial layer of internal organs. The arrayed nanoprobes on the gene chip can breach the mucous layer and extract the transcript mRNAs from internal tissues without the need for tissue biopsy, enabling a bleeding-free in vivo spatial transcriptomic profiling. A pressure sensor is also integrated for controlled application of the nanoprobes in a regular endoscopic surgery, providing the feedback to further minimize tissue damages. In our demonstration with a gastrointestinal (GI) endoscopy system, we successfully obtained multiplexed transcriptomic landscapes of the areas of interest in the intestinal tract of rodent animals or human subjects, which were acquired by a very brief surgical operation on the order of 5 minutes and subsequent off-line informatic analysis. In a clinical study involving 17 human subjects (15 Crohn’s disease (CD) patients, 2 healthy subjects) receiving 47 ENDO-Genome endoscopic examinations, none of the patient develops any complication afterwards. The live spatial transcriptomics provides direct *in vivo* pictures of the heterogenous spatial transcriptional programs underlying CD pathological response at different GI locations, revealing distinct ileal phenotypes. This is manifested by unique microscale scattering of CD inflammation gene clusters, along with the discovery of a tissue-specific cooperative mechanism between CD inflammation and RNA methylation regulations at single- or multi-cell scales.

## Results

### System design

The ENDO-Genome technology relies on the integration of a nano-structured biochip with clinical endoscopic platform that expands its function to transcriptomic profiling of RNA transcripts directly from internal human organs. Meanwhile, the capability for structural and histological examination remains unaffected. The biochip is functionalized for high-throughput extraction of targeted RNA transcripts from internal organs via a “touch & go” operation, which is guided by a typical endoscopic operation via the surgical biopsy port. The integration of a nanoprobe biosensor chip to an endoscope provides a multifunctional instrument to examine transcriptomic regulation in live human tissues in the context of their original in vivo environment (**Figure 1**). When guided to the desired location of an internal organ (e.g. colon), the nano-structured biochip works as a novel biointerface to access the native live tissue environment that harbours rich information related to physiology or pathology of human organs. Particularly for live transcriptomic analysis, RNA transcripts can be in-situ extracted from the organ surfaces with subcellular resolution without damaging local tissues in a bleeding-free format. The biochip is then subjected to off-line informatic analysis to derive the spatial transcriptomic profiles of the endoscopic sampling sites of the examined internal organ, providing diagnostic or therapeutic evaluation.

**Figure 1.**
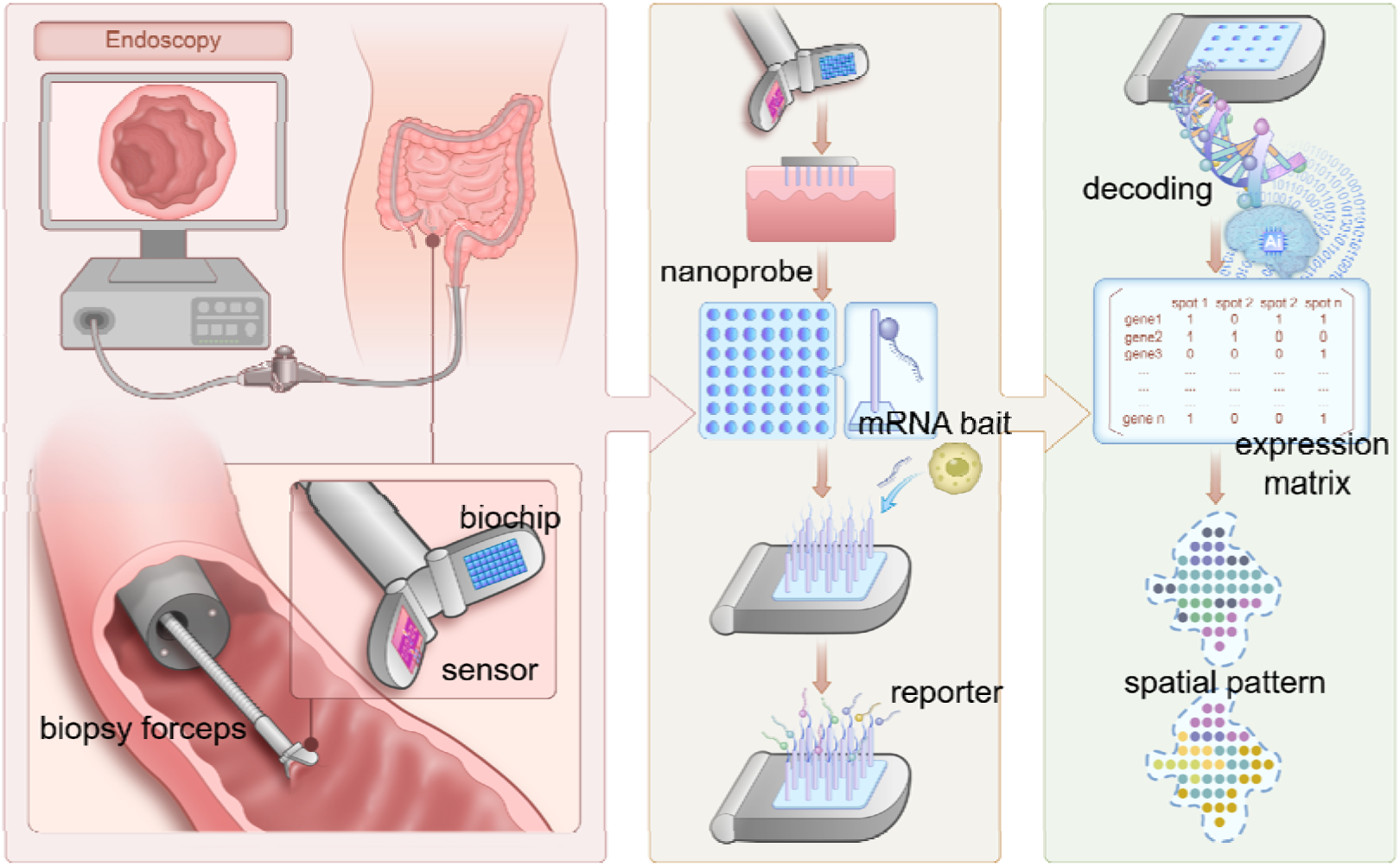
Design of the ENDO-Genome system. The ENDO-Genome system integrates a nano-structured biochip and a mechanical sensor with an endoscopic platform to enable “Touch & Go” extraction of RNA transcripts directly from internal organs on human under precise control by standard endoscopic procedures, creating an in-body spatial transcriptomic technique capable of taking snapshots of in vivo transcriptomic regulation in the living tissue of tis normal or diseased habitat without the need for tissue biopsy, voiding any bleeding risks. This advancement greatly improves the temporal resolution and efficiency to capture the in vivo dynamic and volatile genetic program subject to different post-transcriptional regulations in a cost-effective and clinically accessible way.

In this proof-of-concept study, the ENDO-Genome system is demonstrated with gastrointestinal endoscopy. The functional biopsy port is used for biochip integration and organ targeted in-body sampling. The nanostructured biochip was fabricated from a wafer-scale production involving deep reactive-ion etching (DRIE) process, (**Supplementary Figure S1**). The resulted nanoprobes were measured at 10 µm in height and ∼800 nm in diameter and were arranged in an array with 4 µm spacing distance (**Figure 2a**). The nanoprobes were subsequently crosslinked with polyT DNA sequences for capturing mRNAs Laser cutting was used to fabricate biochips with a customized size (1.6×1.8 mm^2^) adaptable to a biopsy forceps, which is directly loaded to the colonoscope tube and can work with other functional components. Dental cement was used as the adhesive for biochip assembly onto the biopsy forceps to ensure biocompatibility (**Figure 2a, b**). The biochip-loaded forceps is then used for RNA extraction in a complex intestinal environment. The biochip integration does not interfere or compromise the endoscopic operations. The flexible construction of the colonoscope tube ensures precise access of intestine tissues at desired angles under the guidance by the auxiliary imaging system, as we demonstrated in a 3D-printed intestine emulation (**Figure 2c**) or reconstructed *ex vivo* porcine intestine model (**Figure 2d**). The biochip can be navigated to reach all the locations for RNA extraction throughout the gastrointestinal system, including rectum, ascending colon, transverse colon, descending colon, etc.

**Figure 2.**
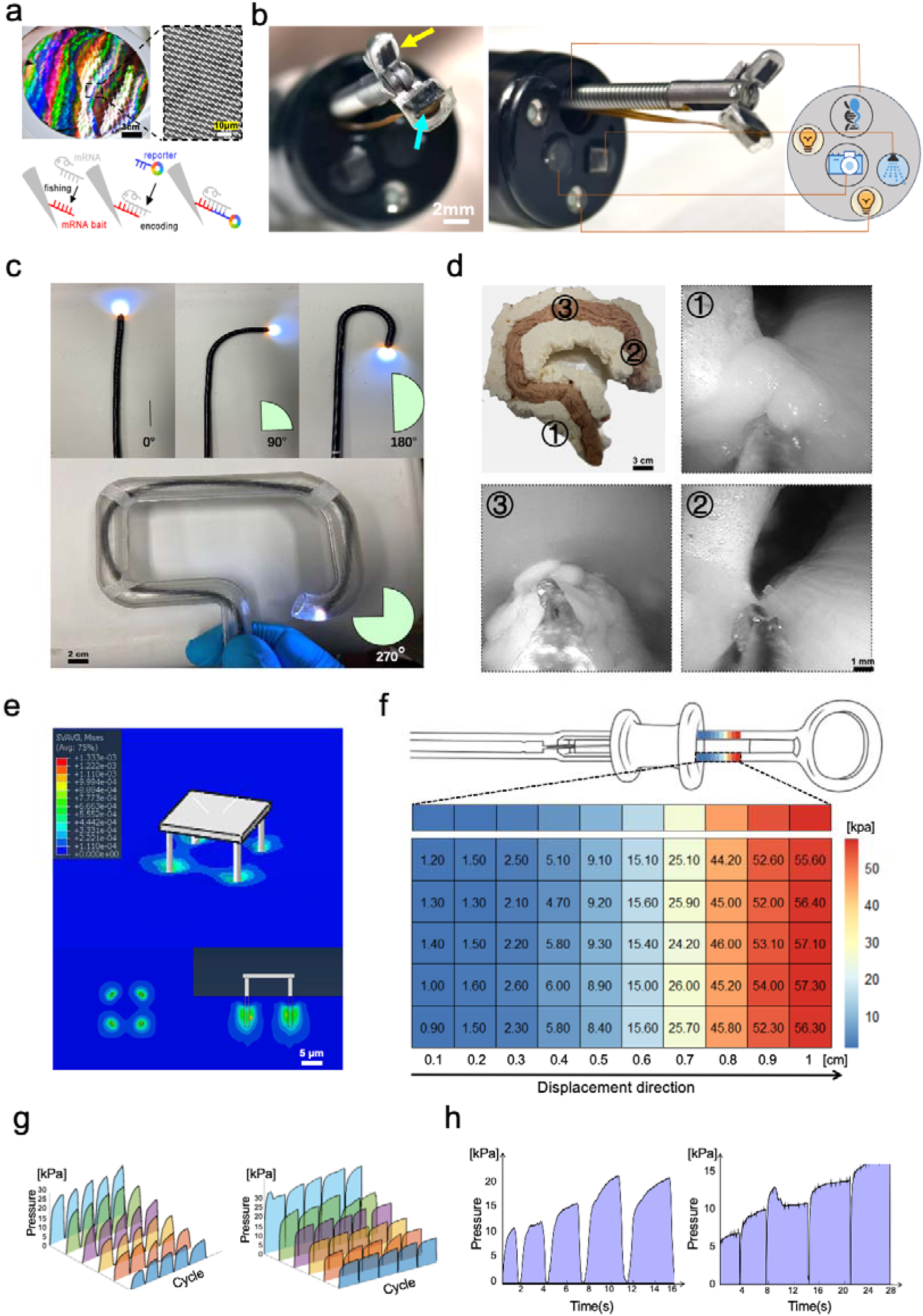
Instrumentation and techical characterization of the ENDO-Genome system. **(a)** Fabrication and functionalization of the nanostructured biochip assembled to the ENDO-Genome system. Top: optical and SEM images of the silicon-based biochip with arrayed nanoneedles from wafer-size fabrication. Scale bar, 3 cm (top left); 10 μm (top right). Bottom: schematic showing the biochemical functionalization of the biochip for molecular fishing of mRNAs from tissues. Bait sequences (polyT) were crosslinked to the nanoneedle for specific capturing targeted mRNAs; fluorescence-encoded reporter sequences then bind to mRNAs for decoding and transcriptomic analysis. (**b**) Integration of the END-genome system, which includes a biochip, a mechanical sensor on a clinical endoscopic platform along with other functional units: biopsy, flushing, and live monitoring. Scale bar 2 mm. (**c**) Characterization of the flexibility of the ENDO-Genome system in a 3D-printed model. Scale bar, 2 cm. **(d)** *Ex vivo* demonstration of the ENDO-Genome sampling process at different intestinal locations (1, rectum; 2, transverse colon; 3, ascending colon) in a porcine intestinal model. Scale bar 3 cm (overall); 1 mm, surgical snapshots in the intestinal tract showing the sampling accessibility. **(e)** Simulation showing the sampling pressure at the interface between the nanoprobes and human tissue, assuming a Young’s module of 3.6 kPa. Scale bar, 5 μm. **(f)** Heatmap showing the calibration of the clamping pressure via a biopsy forceps, which is controlled via the manipulation handle. The variation of applied pressure corresponding to different displacements is shown over a ∼1 cm displacement range. **(g)** Characterization of the dynamics and reliability of the clamp sampling using ENDO-Genome. Multiple cycles of clamp sampling on *ex vivo* tissue in instant (left) and continuous (right) operation modes were shown. **(h)** The temporal dynamics of instant (left) and continuous (right) operation modes of typical ENDO-Genome assays.

### Mechanical sensor for closed-loop in-body transcript sampling

While the EDNO-genome system establishes a rapid and adaptable subcellular interaction between the nano-structured biochip and human tissue in the complex native live-organ environment, the interactive force was controlled by a feedback-clamping system using a highly sensitive thin-film sensor, that was fitted to the 2nd arm of the biopsy forceps opposing the ENDO-Genome biochip to transmit the clamping force data (**Figure 2b**), thus providing delicate surgical control for in-body transcriptomic sampling in the process of a regular endoscopic surgery (**Figure 2e-h**). When interfaced with the intestinal tissue, the EDNO-genome biochip was used to access the cells beyond the intestinal mucosa. By simulation, it is estimated that a single nanoprobe can apply a pressure on the tissue at kPa level, given their microscale interfacing with the tissue, which was calculated by assuming a Young’s modulus of 3.6 kPa for colon tissue ^24^ (**Figure 2e**). At this pressure level, the compressive impact on the tissue is gentle and efficient, reducing the risk of tissue damage. The mechanical sensor ensures safe and reproducible transcript sampling without damaging the tissues, avoiding the risk of intestinal perforation. This is achieved by adjusting the clamping pressure applied through the operation handle with a reference to the continuous and real-time sensor feedback (**Figure 2f**). In an ex vivo sampling calibration on porcine intestinal specimens, the pressure applied at the biochip-tissue interface was typically below 60kPa. The controllable clamping was reliably repeated (**Figure 2g**). If a sustained clamping was applied, a plateaued pressure reading can be readily observable (**Figure 2h**). The ENDO-Genome enables a safe, manageable, and interactive closed-loop in-body transcript sampling process by combining endoscopy clinician expertise with the sensor feedback system.

### Bleeding-free in-body RNA extraction

To evaluate the safe operation of the ENDO-Genome system for in-body profiling human internal organs, the nano-structured biochip-tissue interfacing was first characterized using ex vivo human intestinal tissues. When ∼5kPa pressure was applied to the tissue via a chip, SEM and histology examination showed successful penetration of the superficial layer of the examined tissue by the nanoprobes (**Figure 3a, b**). Beyond the interfacing depth, the cell morphology and tissue integrity were not affected (**Figure 3b**). The silicone based nanoprobes showed excellent biocompatibility with human cells even after prolonged puncturing and incubation procedures (**Supplementary Figure S2**).

**Figure 3.**
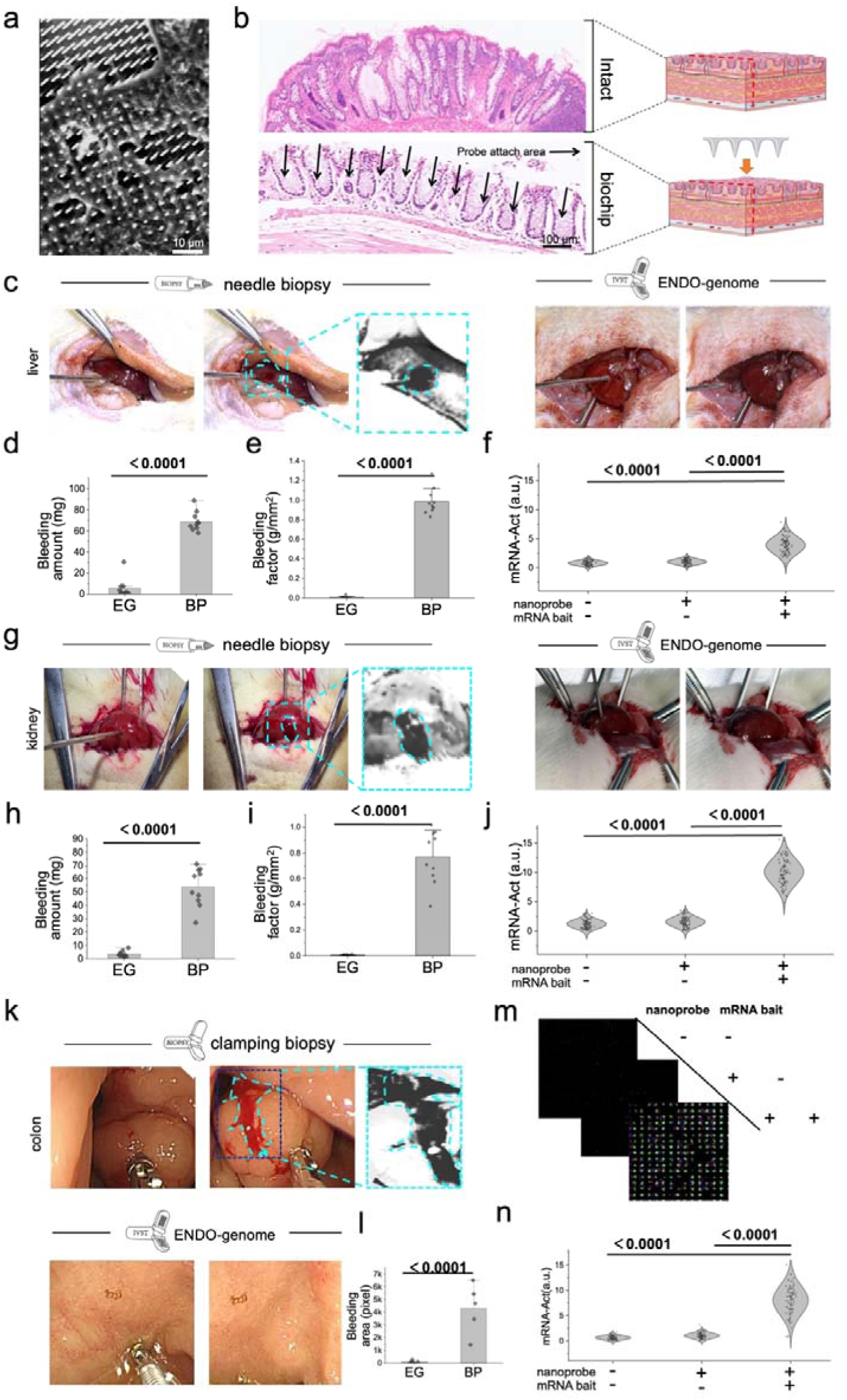
Characterization of bleeding-free in-body RNA extraction using the ENDO-Genome system. (**a**) Scanning electron microscopy (SEM) images showing the biochip and tissue interface. Scale bar, 10 μm. (**b**) Hematoxylin and eosin (H&E) staining of human colon tissues showing the biochip interfacing trace (arrow) after an ENDO-Genome operation. Scale bar, 100 µm. Intact colon tissue was used as the control (top). (**c**) Comparison of bleeding induced by an ENDO-Genome (right) or a typical biopsy needle (left) operation in rat liver. Surface bleeding was outlined and enlarged in the BP demonstration. (**d, e**) Quantification of bleeding amount (**d**) and bleeding factor (**e**) from rat liver after an ENDO-Genome (EG) or a typical biopsy needle (BP) operation. **(f)** Quantification of mRNA (β-actin) extraction from rat liver by an ENDO-Genome operation. (**g**) Comparison of bleeding induced by an ENDO-Genome (right) or a typical biopsy needle (left) operation in rat kidney. Surface bleeding was outlined and enlarged in the BP demonstration. (**h, i**) Quantification of bleeding amount (**h**) and bleeding factor (**i**) from rat kidney after an ENDO-Genome (EG) or a typical biopsy needle (BP) operation. **(j)** Quantification of mRNA (β-actin) extraction from rat kidney by an ENDO-Genome operation. (**k**) Comparison of bleeding induced by an ENDO-Genome (bottom) or a typical clamping biopsy (left) operation in human colonoscopy examination. Surface bleeding was outlined and enlarged. **(l)** Quantification of the bleeding area of human colon tissue treated by EG or clamping biopsy (BP). **(m)** Confocal images showing successful live extraction of mRNA (β-actin) from human colon tissues. **(n)** Quantification of β-actin mRNA signal associated with individual nanoprobes on a biochip. For panel **d**, **e**, **h**, **i**, **l**, data are presented as the mean ± SD, n = 10, analyzed by two-tailed unpaired t-test. For panel **f**, **j**, **m**, **n**, the comparison groups include blank biochip [no nanoprobe (-), no mRNA bait (-)], non-functional biochip [with nanoprobe (+), no mRNA bait (-)] and functional biochip [with nanoprobe (+), with mRNA bait (+)]. >100 data points were collected from 3 biological replicates and analyzed by Mann-Whitney test.

For transcriptomic profiling human internal organs, the ENDO-Genome biochip enables a minimum-invasive access of the native organ environments for direct transcript extraction without the need for tissue biopsy. Therefore, the surgical procedure can be completed in a bleeding-free manner. This is especially important if the technique is applied towards highly vascularized organs, such as liver, kidneys, etc ^25–27^. This feature is then evaluated in rodents by pressurized biochip interfacing with the animal’s liver or kidney (**Figure 3c, g**). A “touch & go” operation on the liver or kidney surface did not cause any bleeding in all examined animals, which would otherwise be much more serious if a traditional percutaneous sampling was performed by using a biopsy needle. Notably, the sampling coverage by the ENDO-Genome biochip was more than 10 folds higher than the tip of the biopsy needle (radius 0.5 mm *vs* 0.15 mm), but measured bleeding was less than 1/12^th^ of a traditional needle biopsy (**Figure 3d, h**). The bleeding factor (weight of operation-induced bleeding divided by contact area) showed more than 100-fold reduction for ENDO-Genome operation in both liver and kidney (**Figure 3e, i**). Successful extraction of RNA transcripts (for β-actin mRNAs) were also achieved in these organs (**Figure 3f, j**).

After these validation in rodent models, the ENDO-Genome assisted bleeding-free profiling technique was further tested in human subjects during a normal gastrointestinal examination. A regular biopsy procedure causes substantial local bleeding in most cases (**Figure 3k**). On the other hand, the ENDO-Genome biochip causes no bleeding in the clamping extraction of transcripts from different sites along the GI system (**Figure 3k**, **Supplementary Movie S1**). This is a direct demonstration for the preservation of in vivo tissue integrity in ENDO-Genome examination, reducing the risk of surgical complications like inflammation and potential infection ^28, 29^. The targeted mRNA transcripts were reliably extracted by the brief interfacing between the nanoprobes and colon tissue in a “touch & go” format. In this simple demonstration, the ENDO-Genome biochip was used to extract β-actin mRNAs from the colon tissue, which was visualized by an off-line imaging-based analytical process after the endoscopic examination (**Figure 3l, m**). The ENDO-Genome bypasses the biopsy procedure by a direct access of the native physiology environments in human bodies, which not only enables a minimum-invasive bleeding-free operation, but also minimizes tissue handling to reduce potential artefacts for accurate capturing of dynamic transcriptomic events.

### Multiplexed in vivo transcriptomics

To demonstrate the potential of the ENDO-Genome for multiplexed in vivo transcriptomic profiling, the technique was firstly assessed in a mice colorectal cancer model by using with a veterinary endoscope system (**Supplementary Figure S3**). The polyp or tumor-like area in the mouse colon was located and sampled, and its transcriptomic profile of 24 RNA methylation related genes were derived across different animals. For multiplexed decoding of the RNA transcripts captured by a ENDO-Genome biochip, we used the Spectrum-CODE^@^ system in the microscopic analysis of an ENDO-Genome biochip, in this animal experiment and later clinical studys (**Supplementary Figure S4**). The results showed a correlation between RNA methylation and tumor pathological progresses in the rodent model (**Supplementary Figure S5**) ^30^. Besides transcriptomic expression profiles, ENDO-Genome also revealed substantial spatial heterogeneity of the examined mRNA transcripts, suggesting it’s great promise for a diagnostic tool using advanced in vivo spatial transcriptomics.

To fully demonstrate the potential of ENDO-Genome for clinical usage, ENDO-Genome procedures were performed at multiple geographic locations in the gastrointestinal system to facilitate the diagnosis and monitoring of Crohn’s disease (CD), which is typically challenging and involves comprehensive approaches combining clinical evaluation, laboratory testing, imaging, and continuous patient assessment^31^. Post-procedural complications were also monitored in the patients. Specifically, the ENDO-Genome biochip was configured to profile 55 mRNA transcripts (**Supplementary Table S1 & S2**), including RNA methylation and inflammation related genes, showing a potential in the diagnostic assessment of Crohn’s disease.

The ENDO-Genome examination was performed on 17 human subjects (15 CD patients and 2 healthy individuals, **Supplementary Table S3**). The 15 CD patients were categorized into “active” or “remission” subgroups by clinical evaluations. Because of the non-destructive advantage, for each subject, multiple “touch & go” RNA extractions were carried out as necessary (determined by the clinician) at different geographical locations along the gastrointestinal system, including cecum, rectum, stomach, ascending colon, transverse colon, descending colon, sigmoid colon, and ileum (**Figure 4a, b**). Accordingly, a total of 47 ENDO-Genome assays were performed. None of the in-body sampling of RNA transcripts caused any bleeding in the patients. The procedure was closely monitored via the regular endoscopic imaging (**Figure 4b)**, as representatively demonstrated by the operation at the ascending, transverse or descending colon (**Supplementary Movie S2**).

**Figure 4.**
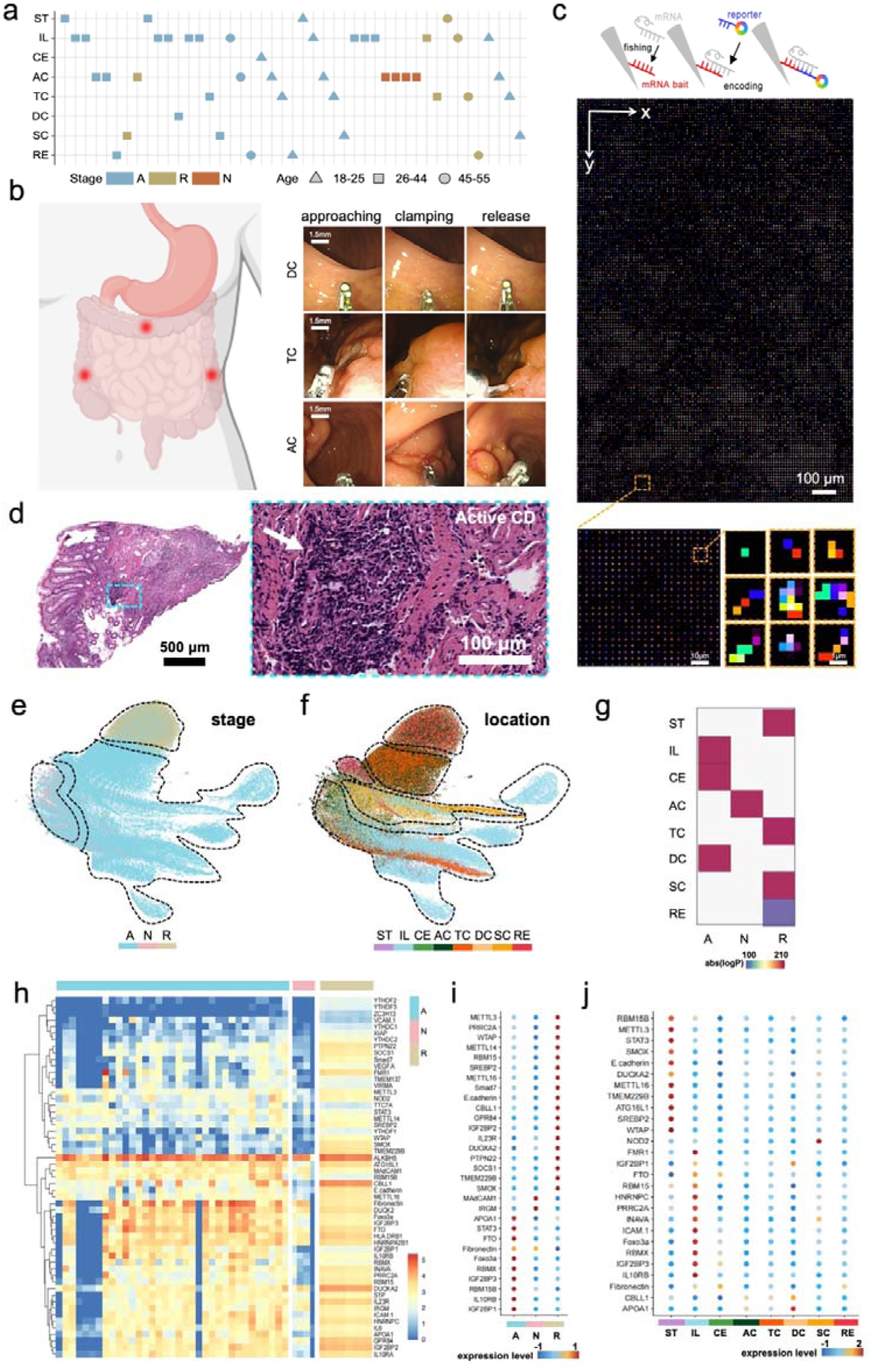
In-body quasi-single-cell transcriptomic analysis in patients with Crohn’s disease. **(a)** Summary of the clinical study information: patient age (circle, 18-25; triangle, 26-44; square, 45-55); disease status (bule, active (A); brown, remission (R); and red, healthy (N)); ENDO-Genome sampling location in gastrointestinal (GI) tract, including cecum (CE), rectum (RE), stomach (ST), ascending colon (AC), transverse colon (TC), descending colon (DC), sigmoid colon (SC), and ileum (IL). **(b)** Representative colonoscopy screenshots showing the ENDO-Genome procedures (approaching, clamping, and release) at different sites (DC, TC, AC) in the intestinal tract. **(c)** Raw imaging data showing the analytical expression of different mRNAs extracted by an ENDO-Genome biochip. Top: Schematic showing the chemical functionalization for molecular fishing of mRNAs. Bottom: enlarged view of the outline regions showing the spectrum-CODE signal associated with each nanoprobes revealing the expression of different mRNAs. **(d)** HE images of intestinal tissues showing the histological CD pathology (indicated by arrow). The outlined region is enlarged on the right. **(e, f)** UMAP embedding illustration of data clustering according to CD status (**e**) or EDNO-Genome sampling locations (**f**). Over 140,000 qSC-spots were derived from over 2000000 nanoprobes using 4×4 binning for analysis. **(g)** Hypergeometric test revealing the significant association between the clustering labeled by CD status and the clustering results based on the geographic GI sampling. Matrices with p<0.05 are presented. The color indexing is represented by log(P-value); darker indicates more significant association. **(h)** Heatmaps showing expression patterns of the 55 mRNA transcripts across the 17 human subjects (15 CD patients, 2 healthy subjects). The rows represent different mRNA transcripts; the columns represent 47 sampling using different biochips (35 active, A; 8 remission, R; 4 healthy, N). **(i, j)** Bubble plot of differential expression analysis illustrating the dominant transcriptomic expression in different CD status or in different GI locations. For **i** and **j**, the analysis was conducted using Seurat; the genes were selected based on a threshold of |log(fold change)|>1 and p<0.05 (determined by Wilcoxon rank-sum test); the color indexing indicates the expression level.

Upon completion of the ENDO-Genome examination, the biochips were disassembled from the biopsy forceps and analyzed using an imaging-based protocol to derive the transcriptomic profiles for the sampled GI sites. In this clinical study, the nanoprobes on the biochip were functionalized with polyT DNA probes for in vivo “molecular fishing” of mRNA transcripts. The panel of 55 mRNA transcripts were carefully selected to be highly correlated with the pathogenesis or progress of CD (**Supplementary Table S2**). By using the spectrum-CODE^@^ platform, functional encoding and decoding of the transcriptomic targets were achieved using a 7-channel fluorescent system ^32^. For every nanoprobes, the captured mRNAs were visualized and quantified with an abundance-indicative expression data and the associated spatial coordinates (**Figure 4c**). As the core advantage of the ENDO-Genome system, the preservation of cellular spatial information associated with the acquired transcriptomic profiles is directly reflected by the spatial distribution of the nanoprobes with their implied registration to the cells within the native in vivo environment, which enables analysis of the susceptibility and spatial reorganization of pathological biomarkers. Particularly for CD, this kind of the disease-indicative information can be observable in the histological analysis for the colon tissue extracted from patients. For example, the samples in the active group showed pronounced accumulation of inflammations and abnormalities (**Figure 4d**). ENDO-Genome further provides extra dimensions of information in association with and beyond these pathological observations by enabling spatial transcriptomic analysis.

In this study, the transcriptomic data acquired from the 47 biochips were pooled together for informatic analysis. The UMAP embedding displays sample clustering based on transcript expressions, showing clear separations for the data points from the active, remission, or normal (healthy) groups (**Figure 4e**). These data suggest that the expression profile of the selected mRNAs could be a molecular phenotype related to the disease progression for CD patients. It was noticed that sampling at different GI locations also forms patterned distributions in the UMAP space (**Figure 4f**). Data from ascending colon (AC) and descending colon (DC) exhibited highly similar transcriptomic features. In contrast, data from sampling at ileum showed well-separated distribution from other locations, implying its unique heterogeneity in CD (**Figure 4f**).

For the pooled data, hypergeometric test revealed that there is a significant association between the clusters labeled by CD progression (A, active; N, normal; R, remission) and the clustering results based on the geographic GI sampling. Particularly, the active mRNA cluster was observed to be particularly enriched by nanoprobes sampling in the and ileum (IL), cecum (CE) and DC, while the remission mRNA clusters was observed to be particularly enriched by nanoprobes sampling in transverse colon (TC), sigmoid colon (SC) and stomach (ST) (**Figure 4g**), suggesting the existence of a geographic heterogeneity of disease proregression in the CD patients, which echoes the known features of CD for its wide affecting coverage and segmental inflammation in the intestinal tract ^33^. Such heterogeneity can be directly observed in the heatmap transcriptomic patterns across the 47 assays from 17 subjects. While the transcriptomic profiles derived from different individual human subject also showed some variations (**Supplementary Figure S6**), on average (across all nanoprobes of a biochip), the mRNA transcripts were grouped into two major clusters (**Figure 4h**), one of which is enriched with several genes related to CD inflammation. For example, IL10RB, FTO, Foxo3a and IGF2BP3 were found to be upregulated in active CD groups. This observation is well in line with the recent literatures regarding the genes associated with CD susceptibility and prognosis ^34–36^. Statistically, different gene subsets were found to be dominantly regulated in samples of different CD statuses (active, remission, normal) (**Figure 4i**). Sampling at particular GI locations (e.g. IL, ST) also showed unique regulation by specific genes (**Figure 4j**). Interestingly, we found substantial overlapping between the IL-dominant and the active-dominant genes. For example, the expression of IL10RB, IFG2BP3, RBMX, FTO and Foxo3a were all identified to be significantly upregulated in active CD patients or in the ileum samples (**Figure 4j, k**), as quantitatively illustrated by differential expression analysis using bubble plots. It is known that ileum is the most commonly involved intestinal location in CD, although the disease can globally affect any site of the GI tract ^37^, our results directly provide the in vivo transcriptomic evidence confirming the segmental feature in active CD patients, which promise novel insights to CD monitoring.

### Unsupervised clustering for transcriptomic analysis

As the ENDO-Genome biochip interfacing with the internal colon tissue for the “touch & go” RNA extraction, the nanoprobes effectively sampled the touched colon tissue with subcellular resolution. Effectively, the in vivo spatial transcriptomic heterogeneity is preserved by the nanoprobe associated transcriptomic expressions. To explore such information’s association with CD, we then performed unsupervised clustering of mRNA expressions derived from the 140,000 quasi-single-cell spots (qSC-spots, 4×4 binning from >2000000 nanoprobes), which were respectively acquired from 47 sampling studys in the 17 subjects. The analysis identifies 10 qSC-spot clusters (C1–C10) in the pooled mRNA data. To visualize and compare the clustering patterns using UMAP embedding, dimension reduction was performed on the mRNA expression in the multidimensional space formed by the 55 selected mRNA transcripts (**Figure 5a**). For each qSC-spot cluster, the average mRNA expression across all nanoprobes was illustrated in a heatmap to further identify cluster-specific mRNA signatures (**Figure 5b**).

**Figure 5.**
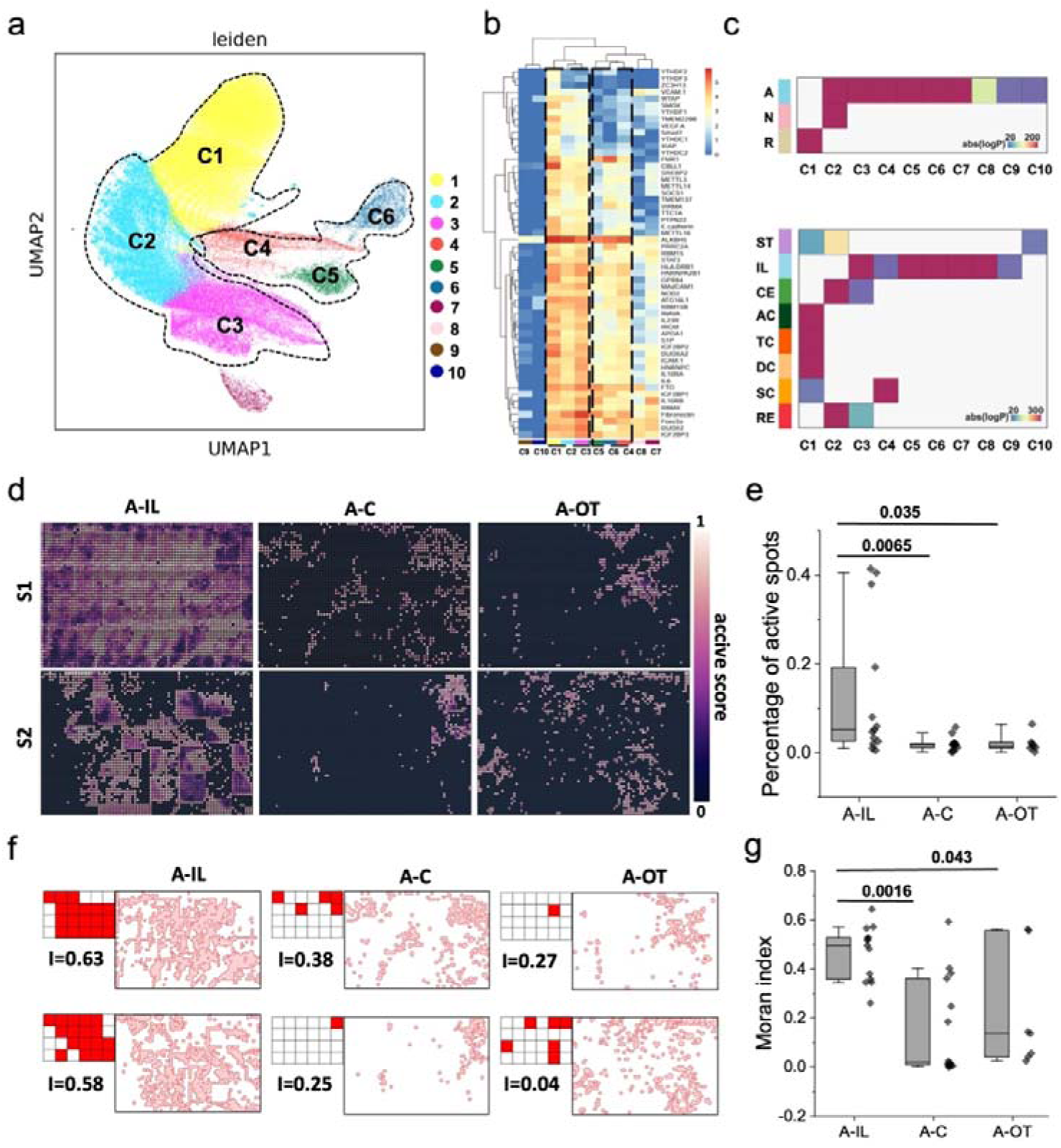
Unsupervised clustering for spatial transcriptomics analysis. **(a)** UMAP embedding illustration of the unsupervised data clustering analyzed by Leiden algorithm. The data set contains 140,000 qSC-spots derived from >2000000 nanoprobes using 4×4 binning. Each point represents the two-dimensional projection of an individual qSC-spot associated with expression of 55 mRNAs, illustrating the intrinsic coherence based on gene expressions. **(b)** Heatmap of averaged expression of individual mRNAs across the10 qSC-spot clusters derived by unsupervised clustering. **(c)** Enrichment analysis using the hypergeometric test showing the significant association between qSC-spot clusters CD statues (top) or GI sampling locations (bottom). Matrices with p<0.05 are presented. The color indexing is represented by log(P-value); darker indicates more significant association. **(d)** Representative spatial scattering patterning of high CD active score (A-score) in ENDO-Genome sampling at different GI locations in active CD patients (2 were shown, A-IL: active-ileum, A-C: active-colon, A-OT, active-other GI locations). For each qSC-spot, its A-score was calculated by taking a linear combination of its Euclidean distances to the signature vector of each qSC-spot cluster. **(e)** Boxplots showing the quantification of the portion of high A-score qSC-spots of individual sampling at different GI locations in active CD patients. **(f)** Spatial distribution of active spots (A-score>0.7) and the associated Moran’ index in samples of different GI locations in active CD patients. Moran’ s index plot is generated from original spatial scattering plot by binning and binarization. **(g)** Boxplots showing the quantification of the Moran’s index of individual samples from different GI locations in active CD patients. For **e**, **g**, the dots indicate an overlay of the data points, each box indicates the first quartile, median, third quartile, the whisker limit is 1.5 times the interquartile range, p-value is determined by Kruskal-Wallis test.

Enrichment analysis using the hypergeometric test suggested that several qSC-spot clusters (2-7) were observed to have significant association with the “active” CD labeling of the data. Interestingly, a substantial portion of the qSC-spot clusters (3, 5-8) also showed significant association with the transcriptomic sampling at IL (**Figure 5c**). The majority of the two associative qSC-spot clusters overlapped (3, 5-7), indicating IL as the most affected GI geographic location in active CD patients by the transcriptomic profiling. Then, each of the overlapping qSC-spot cluster (3, 5-7) was used to derive a 55-dimension cluster signature vector by taking the average across all associated qSC-spots. The signature vectors are then used to derive a CD activity score (A-score) for individual qSC-spots across an ENDO-Genome chip. For each qSC-spot, its A-score was calculated by taking a linear combination of its Euclidean distances to the signature vectors. Higher A-score indicates more closer expression pattern to the signature vectors, indicating higher level of CD pathology. As representatively shown in **Figure 5d**, the IL tissue in active CD patients showed substantially more qSC-spots of high A-score than other GI tissues in these patients (**Figure 5d, e**). Meanwhile, the high A-score qSC-spots also showed different levels of clustered distribution across an ENDO-Genome chip covering 1.6×1.8 mm^2^ tissue. Such spatial phenotype was quantitively analyzed by using Moran’s index (MIs): higher indices indicate greater clustering of data points of similar features ^38^ (**Figure 5f**). Statistically, the Moran’s indices were significantly higher for IL sampling than other GI locations in the active CD patients (**Figure 5g**), and were also significantly higher than the results derived from sampling in non-active subjects (**Supplementary Figure S7**). These results suggest the great diagnostic potential of transcriptomic profiling of human internal organ by using the ENDO-Genome system for determining the multiplexed transcriptomic alternations in CD patients.

### CD-indicative spatial transcriptomic patterning

Focusing on individual genes, the heatmap of their MIs showed elevated levels for a good portion of the transcriptomic targets in the expression profile derived from the IL of active CD patients, when compared to the sampling at other GI locations (**Figure 6a**). Many of the identified gene, such as Foxa3a, IGFBP3, FTO, and IL10RB, showed substantially clustered distributions (**Figure 6b**), which echoes well with our previous analyses based on the *in vivo* mRNA expression levels (**Figure 4**). Surprisingly, some genes did not show any specific expression (**Figure 4h-j**) but turned out be uniquely regulated in active IL, as indicated by the spatial parameter using MIs. For example, NOD2, the widely reported CD risk gene with differential association in western or asian populations ^36, 39^, was not particularly up- or down-regulated in active CD patients (all eastern asians) or their IL tissues, but was discovered to show some level CD-indicative scattering in the active IL (**Figure 6c**), providing a new perspective for understanding the heterogeneity of CD pathogenesis using spatial transcriptomics. When the gene-specific MIs were pooled together and statistically compared across different in-body sampling locations (ileum, colon, others) in active CD patients, showing significantly higher MIs for the IL sampling versus other GI sites (**Figure 6c**). In particular, the statistic on individual transcripts further highlights the CD-pathology associated phenotypes indicated by the spatial organization of Foxa3a, RBM genes, and IGFBP1/2/3 genes (**Figure 6d**). These results directly demonstrated the clinical promise of ENDO-Genome to provide disease indicative spatial transcriptomic information beyond simple expressions.

**Figure 6.**
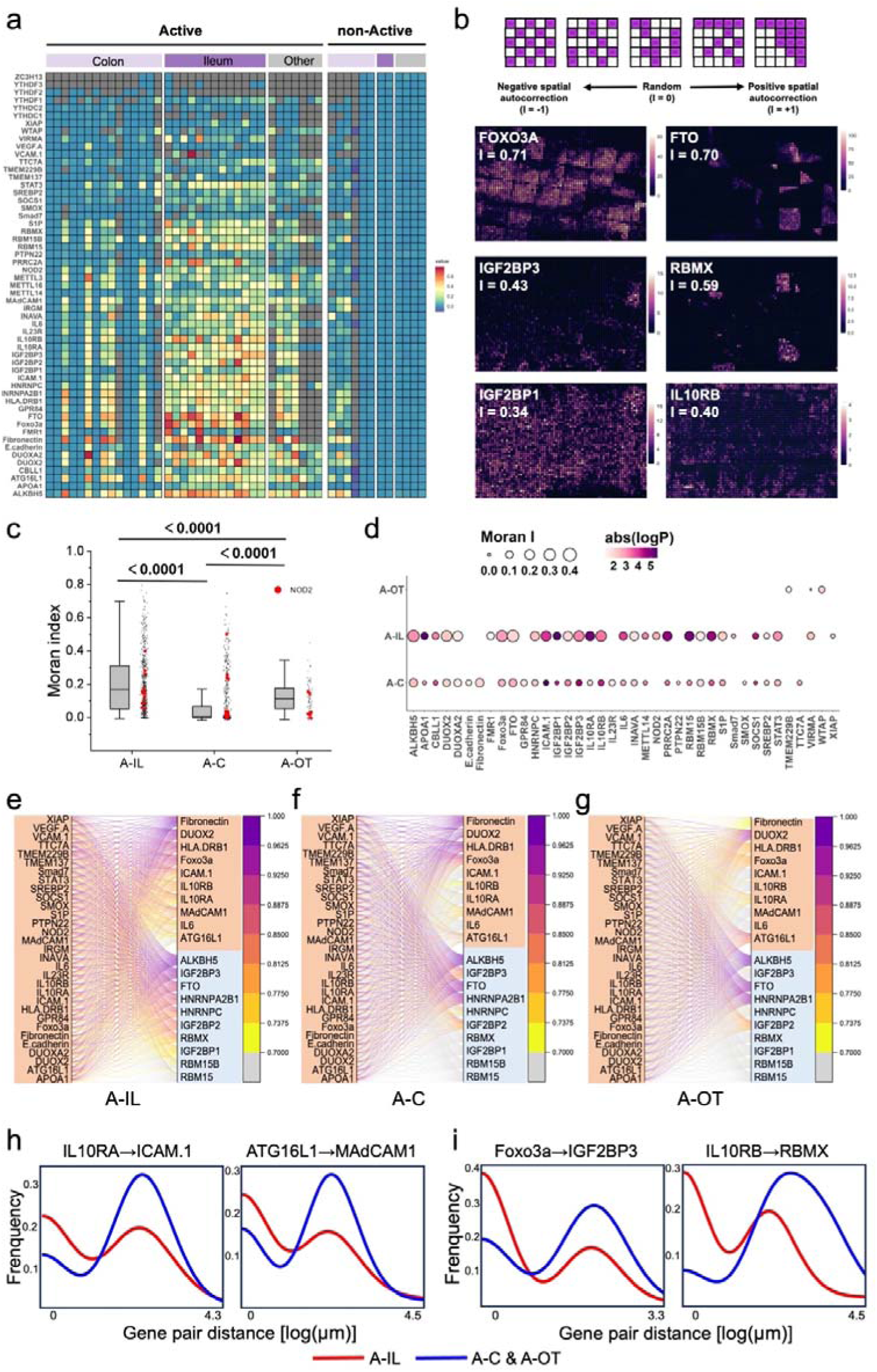
CD-indicative spatial transcriptomic patterning. (**a**) Heatmaps of Moran’s Index for different CD-related genes across all the sampling in human subjects. The samples are categorized by CD status (Active *vs*. non-Active) and GI sampling location (colon, ileum, other). (**b**) The spatial expression scattering of representative genes identified to be highly affected in active ileum, showing different Moran’s indices. (**c**) Boxplots showing the quantification of Moran index of individual genes pooled from all sampling at different GI locations in CD patients (A-IL, active-ileum; A-C, active-colon; A-OT, active-other). The black dots indicate an overlay of the data points, and the red dots indicate an overlay of data point for NOD2 gene; each box indicates the first quartile, median, third quartile of the data, the whisker limit is 1.5 times the interquartile range, p-value is determined by Kruskal-Wallis test. (**d**) Statistical analysis of Moran indices of individual mRNA transcripts showing CD-associated spatial organization of different genes. The color index indicates log(p-value), the dot size indicates averaged Moran index of a gene. Differential dominance of a gene in a category (A-IL, A-C, A-OT; one *vs.* all others) is determined by Wilcoxon rank-sum test with significant p< 0.05. (**e-g**) Analysis of distance-dependent transcriptomic patterning for gene-pairs at single cell scale (<20 μm) in active CD patients. Comparison is made among A-IL (**e**), A-C (**f**) and A-OT (**g**) categories. Mapping from the anchor genes (left column), the spatial distance of the closest expression of the pair gene (right column) was prioritized in the inflammation (brown) or methylation (blue) regulator groups. The associative probability was indexed by a colored line connecting the anchor and the pair genes with a threshold set at 70%. (**h, i**) Histogram of the association distance for representative gene pairs, comparing inflammation-responsive participation of endothelial dysregulation (**h**) or RNA methylation (**i**) related genes in the ileum (red curve) of active CD patients.

In theory, the spatial transcriptomic phenotypes of the healthy or diseased human tissues are linked to the heterogeneous cellular organizations, which could be captured by the patterned regulation of transcript pairs or groups on the scale within or across individual cells ^40, 41^. For ENDO-Genome profile acquired from the human subjects, we then investigated the distance-dependent transcriptomic patterning for gene-pairs on different scales. In the active CD patients, it was found that the expression of CD-related inflammation genes has a high probability (>70%) of accompanied expression of Fibronectin and Foxa3a over a short-distance scale less than 20 μm across different locations of the intestinal tracts (IL, colon, other locations) (**Figure 6e**, **Supplementary Table S4**). Some RNA methylation genes (e.g. FTO, ALKBH5, IGF2BP2) were also observed to be responsive associated with CD inflammation (**Figure 6e-g**). Beyond these geographically common patterns, spatial transcriptomic profile of active IL sampling further showed unique paired gene expressions. Association between CD inflammation to ICAM.1 and MAdCam1 were specifically identified at IL (**Figure 6e, h**), potentially suggesting a unique role of endothelial dysregulation contributing to the alleviated inflammation of IL tissue in CD patients ^42^. On the epigenetic level, specific association of RBMX, IGF2BP3, HNRNPC with CD inflammation genes were uniquely increased in IL than in other GI locations (**Figure 6e, i**), further indicating the distinct responsive participation of RNA methylation in regulating intestinal inflammation over CD progression. Interestingly, the pattern of conditional gene pairing changes over a large-distance scale (>80 µm). The ileal expression of CD inflammation genes is frequently (>70% probability) accompanied by the expression of ZC3H13, YTHDF3 and YTHDC1 (**Supplementary Figure S8**, **Table S5**), suggesting the possible existence of cross-cell epigenetic regulation by the involvement of multiple cell populations ^33, 43^. Altogether, these results provide direct in vivo pictures of the spatial transcriptional programs underlying CD pathological response at different GI locations, showing distinct phenotypes attributed to the IL.

## Discussion

In this study, we develop ENDO-Genome as an advanced spatial transcriptomics technology for live profiling human internal organs with single-cell resolution, thus providing disease-indicative transcriptomic information that is currently inaccessible to existing methods. This is achieved by integrating a nanoarrayed biochip with gastrointestinal endoscopy system to perform minimally invasive in-body RNA extraction for spatially resolved transcriptomic analysis of human internal organs. As part of the invention, the ENDO-Genome system takes advantage of a sequencing-free approach ^44^, which features a “Touch & Go” RNA extraction to enable the massively paralleled acquisition of targeted RNA transcripts without the need for tissue biopsy. In a completely bleeding-free manner, ENDO-Genome operations could be performed at multiple locations of the GI system to resolve the geographic heterogeneity of transcriptomic patterning of an organ level scale. Notably, we also demonstrated very low bleeding risk but excellent mRNA yield in rodent liver and kidney. This is thought to have important clinical implications because conventional biopsy procedures in liver and kidney are usually of high-risk for bleeding and difficult to stop. This ENDO-Genome technique emerges as a safe approach obtain single-cell level transcriptomic information directly from these highly vascularized organs. In the clinical studys involving 15 CD patients, the colon wall of the active subjects was compromised with different levels of CD pathologies, including transmural inflammation, fissures and ulcers, not a single complication case was observed out of a total of 47 ENDO-Genome operations, showcasing the unique safety and gentle deposition of the technology.

On the cellular scale, the biochip approach preserves the cellular spatial information associated with the acquired phenotypic profiles (e.g. mRNA expression), which is directly reflected by spatial distribution of the nanoprobes with intrinsic spatial registration to the cells in examined tissue. This enables analysis of the susceptibility and spatial reorganisation of different pathological biomarkers directly in the tissue being sampled by the ENDO-Genome operation. The transcriptomic quantification of each nanoprobe is analyzed with subcellular resolution, allowing for quasi-single-cell analysis to decipher the tissue-wide spatial transcriptomic heterogeneity. Over the past decade, significant progresses have been made in developing spatial transcriptomic technologies with improved the resolution, sensitivity, and accessibility ^45^. Techniques like Slide-seq, STARmap, and Genomics Visium provide the capability to analyze large tissue sections ^5, 45, 46^. Alternatives based on *in situ* hybridization allow researchers to achieve single-cell resolution, providing more detailed spatial maps of gene expression ^47^. More recently, more affordable and convention solutions are emerging to further push the boundaries of spatial transcriptomics ^48^. There is also a growing trend of integrating spatial transcriptomics with other omics technologies for more comprehensive understanding of cellular heterogeneity in normal or diseased conditions ^49^, with the great promise of benefiting clinical diagnostics and treatment ^50^. However, current usage of spatial transcriptomics is largely untapped for disease management, due to prohibitive cost and technical complexity. This critical void emphasize the unique advantages of the ENDO-Genome by providing a low-cost and affordable spatial transcriptomic analysis on human tissues without the need for histology processing, presenting an opportunity to supplement clinical practices with the most accessible spatial genetic or epigenetic information, which typically undergoes volatile dynamic regulation ^51^, and can be lively captured from the in vivo native habitat by ENDO-Genome. This is particularly useful for bridging the spatial transcriptomics across patient’s tissue or organ with disease diagnosis, drug application, and treatment customization.

From an instrumentation perspective, ENDO-Genome also an innovation towards next-generation endoscopy technology. Endoscopy techniques allow for minimally invasive procedures to visualize and examine of internal organs using a flexible tube equipped with a camera and surgical tools ^52^. For example, colonoscopy plays a crucial role in the early detection, diagnosis, and management of various gastrointestinal conditions: it is the gold standard for screening and detecting colorectal cancer, and is also extensively used for diagnosing conditions like Crohn’s disease and ulcerative colitis by allowing direct visualization of the intestinal lining ^53^. While endoscopy technology continues to evolve, existing endoscopic tools only provide structural examination or some sampling capabilities, but not functional information regarding the subtle changes regarding the molecular or cellular phenotypes in the examined tissues or organs. As we demonstrated technically and clinically, ENDO-Genome provides one type of such missed information, the change of transcriptomic regulations, in the native environment within human body, which is inaccessible by any existing techniques and leads to the pathway towards a next-generation endoscopy system with both structural and functional reporting capabilities.

Clinically, the diagnosis and longitudinal monitoring of CD need a comprehensive approach involving colonoscopy, imaging, laboratory tests and other methods ^54^. More recently, translating gene expression signatures and multiomic data into tangible understanding of CD is believed to be promising to guide disease treatment but is still challenging ^33, 36^. The spatial heterogeneity of gene expression within the CD affecting tissues or the geographical variation along the largescale GI tract is largely unexplored. In most cases, transcriptomic assays are performed on biopsy tissues extracted by local destruction over endoscopic examinations, which may cause intestinal flora disturbance, local bleeding, and even perforation, posing risks to patients. Especially, given the life-long recurrence risks and systemic nature of CD, single-site biopsy is typically insufficient, but multi-site endoscopic biopsy poses substantially increased uncertainty for a patient. The out-of-body tissue processing could further induce unwanted artifacts, which especially important if the volatile and dynamic epigenetic targets are used for diagnostic purpose ^55–57^.

As we demonstrated in this study, the technology advancement brought by ENDO-Genome has enabled the discovery of multi-aspects CD-indicative transcriptomic pathology. These data function as snapshots of the disease that can be mined and integrated with other studies to provide insights into CD heterogeneity at the level of pathogenesis and treatment response ^34^, resolving the multigenic involvement of genetic, immune, gut microbiota and environmental factors ^37^. As a relapsing-remitting form of inflammatory bowel disease, it was not surprising to found that many inflammation genes (e.g. IL10RB, Foxo3a) were significantly affected in active CD patients. When spatial information (e.g. the MIs) are considered at cellular level, the ENDO-Genome profiling in different intestinal tissues also showed unprecedented disease-indicative potential. For example, it was encouraging to discover a unique spatial pattering of IGFBP1, IGFBP2 and IGFBP3’s in the IL of active CD patients (**Figure 6e-g**), as these factors have been identified as genetic risk factors for severe complications associated with CD prognosis ^34^, which was also found to be closely associated with Foxa3-driven pathway modulating inflammatory response ^35^. These connections are well supported by the spatial scattering of extra relevant signaling gene pairs revealed in this study (**Supplementary Figure S9**). Particularly, the in-body “Touch & Go” transcript extraction make it possible to snapshot the in vivo transcriptomic regulation in the living tissue of tis normal or diseased habitat, including the microbes, which is previously inaccessible and greatly improves the temporal resolution and efficiency to capture the dynamic and volatile genetic program subject to different post-transcriptional regulations. Further benefiting from the bleeding-free and biopsy-free ENDO-Genome operation, multi-site examination along different GI locations could be safely and easily performed in one patient using a regular colonoscopy protocol, thus providing the necessary data help explain the intestinal geographic heterogeneity of CD and the differential association risk genes in distinct human populations ^54^.

While the exact mechanisms behind the discoveries made here remains to be carefully explored, the results shed lights on a novel perspective for understanding and monitoring CD pathogenesis and progression. In a pilot analysis of the follow-up with the patients involved in this study, some spatial transcriptomic signatures were preliminarily discovered to be associated with the prognosis of these CD patients (**Supplementary Figure S10**). Potentially, the in vivo spatial transcriptomic technology could further benefit personalized design of treatment strategies for CD patients. For example, the choice of drugs may be guided by targeting different pathways in association with distinct transcriptomic patterning in the patients. Accordingly, the use of ENDO-Genome promises a solution to resolve the diagnostic and therapeutic insufficiency with the latest advance in spatial transcriptomics, potentially leading to improved treatment outcomes, increased quality of life, and reduced economic burden for patients.

Beyond the demonstration with colonoscopy in this study, the ENDO-Genome is readily extendable to other endoscopic system (e.g. bronchoscopy, cystoscopy, laparoscopy, etc.) or surgical tools (biopsy needles) for examining different organ systems with unprecedented convenience and cost-effectiveness, which is expected to greatly grow the accessibility and clinical integration of spatial transcriptomic technology.

## Methods

### Animals

All animal procedures were approved by the Animal Ethical Committee of City University of Hong Kong (CityU) and Department of Health of Government of Hong Kong SAR (DH/HT&A/8/2/5/Pt.7). Female C57BL/6 mice (6-8 weeks) and adult male rats (Sprague–Dawley, 200–220g) were acquired from the Laboratory Animal Research Unit (LARU) of CityU.

### Clinical study

All procedures involving human samples were performed in accordance with the ethical standards of the institutional and national research committee under the 1964 Helsinki Declaration and its later amendments or comparable ethical standards. Ethical approval (2024ZSLYEC-304) with informed consent was obtained at the Sixth Affiliated Hospital of Sun Yat-sen University.

17 Human subjects were recruited at the Sixth Affiliated Hospital of Sun Yat-sen University and included 15 CD patients and 2 healthy subjects. The researchers have received necessary training about the research program, and have sufficient understandings of the research. Dedicated quality control personnel check the clinical execution to ensure that all the study conditions meet the research and clinical requirements of the program. Throughout the study, the researchers carry out the clinical operation according to the standard principle and procedures as outlined in the and the approved research protocol. The corresponding project documents were achieved and preserved by the quality control personnel. The quality assurance department of the clinical research unit (Sixth Affiliated Hospital of Sun Yat-sen University) audits the clinical research.

The nature, objectives, procedures, expectations, timing, potential risks and benefits, and any discomfort that may arise from the operation have been explained in detail to each subject with informed consent. Each subject was informed that his/her participation in the study was voluntary, that he/she could withdraw at any time, and that withdrawal of informed consent would not affect his/her subsequent treatment.

### Fabrication and functionalization of nanostructured biochips

Biochips with silicon nanoprobes were prepared using a top-down MEMS approach. An array of dots (1 μm in diameter, spaced by 4 μm center-to-center) were produced by photolithography on a silicon wafer. Next, deep reactive ion etching (DRIE) was performed to fabricate an array of micropillars (∼10 μm height), which were further processed by thermal wet oxidization and chemical etching to reduce the diameter to around 0.8 μm. The silicon wafer with the nanoprobes were then cut into square chips (1.6×1.8 mm^2^) by using a UV laser.

For functionalization, the nanoprobe biochips were processed with piranha solution (v/v = 3:1, 98% H_2_SO_4_:27.5% H_2_O_2_) at 90 □ for 90 min to produce surface hydroxy, and subsequently rinsed with deionized water, methanol, methanol/dichloride methane (DCM) mixture (v/v = 3:1). The chips were immersed in DCM solution containing 3% (v/v) (3-aminopropyl)triethoxysilane (APTES) for 3 hours to render amine groups on the nanoprobe surface. Afterwards, the chips were rinsed by ethanol, isopropyl alcohol and deionized water. The amine groups were then activated by glutaraldehyde (15%, v/v) treatment for 2 hours and then crosslinked with the functional proteins, streptavidin (SA, for mRNA chip, 10 μg/ml), for 2 hours. The chips were then placed in 1% BSA solution (in PBS) with the addition of 0.1% triton X-100 for 5 hours to block the unreacted groups. The biochips were further reacted with biotinylated polyT sequences (100 nM) for 1 hour, followed by incubation of biotin (0.1 mg/ml) for another 2 hours to block the unreacted binding sites on the SA proteins. All the reactions were performed at room temperature unless specifically mentioned.

### Instrumentation of the ENDO-Genome devices

The ENDO-Genome device is an integrative assembly of a functionalized biochip, a piezoresistive pressure sensor, and a set of biopsy forceps (Vedkang, Jiangsu, China) to a gastrointestinal (GI) endoscope system (Olympus). To assemble the biochip to the clamping hand of the biopsy forceps, mineral trioxide aggregate (MTA, Guanya Ltd., Beijing, China) was used. First, 0.15g powder component was added to 0.1 ml liquid component to prepare a viscous MTA mixture. The MTA was dispensed to the surface to one clamping hand and the bottom of the biochip (1.6 × 1.8mm^2^), which were assembled together after the solidification of the MTA adhesive at room temperature (25°C) for 2 minutes. On the opposite hand of the clamping forceps, the pressure sensor was assembled, with its electric cable encapsulated with insulation tape (3DM Ltd.) and winded through an aperture (5mm diameter) towards to the operation handle. The biopsy forceps assembled with a biochip and a pressure sensor were inserted into the functional port of a GI endoscope for accessing the intestinal tissues.

### Establishment of ex vivo intestinal model

As a simple and artificial intestinal model, a tubular structure was fabricated by using a 3D printer (Objet260 Connex3, Stratasys, Eden Prairie). Each layer has a thickness of 2 mm. To maintain the geometric fidelity for printing, the internal cavities were filled with wax supporting material (SUP706). After 3D-printing, the model was immersed in an acetone bath (≥99.5%, Sigma-Aldrich) for 10 minutes to remove the supporting material before usage.

Another *ex vivo* intestine model was established by using fresh porcine intestine, which was anatomically stabilized by casted polyurethane foam. Fresh porcine intestine was folded into an anatomically relevant conformation, recapitulating human *in vivo* spatial intestinal structures. Polyurethane foam (3DM Ltd.) was sprayed to the surrounding space to encapsulate the intestinal segments. The polyurethane was allowed to cure at 25°C for 30 minutes, achieving rigid and stable fixing the intestine tissue, and also creating an opaque sealing exterior to test the endoscopic operations.

### Quantification of bleeding risk of ENDO-Genome

To analyze the bleeding risks in transcriptomic sampling by ENDO-Genome in liver or kidney, rats (Sprague–Dawley, 200–220g) were gas anesthetized with Isoflurane. Then biopsy needle tissue extraction or ENDO-Genome transcript sampling were tested at the exposed liver or kidney of the animals. The induced bleeding was first quantified by measuring the amount of blood after an operation. Before a test, the hemostatic cotton was weighed; after a test, the same cotton was applied to the operation site for blood collection. The induced bleeding was quantified by measuring the weight gain of the hemostatic cotton:

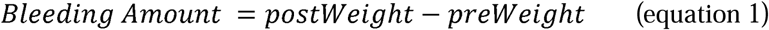

Furthermore, the parameter, “bleeding factor” was defined as the ratio of bleeding amount per contact area of the sampling interface, which was calculated by:

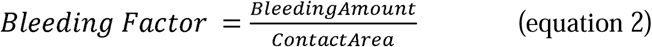

### Multiplexed encoding and decoding for on-chip transcriptomic analysis

For multiplexed encoding and decoding of ENDO-Genome extracted RNA transcripts, on-chip hybridization with targeted RNA transcripts was performed by addition of the DNA-encoded reporter sequences, which were also fluorescently labeled by a specific Spectrum-CODE^@^ beads for visualization in later confocal microscopy. The Spectrum-CODE beads were commercially acquired (300-400 nm in diameter, ANRim Biotech). A 7-channel Spectrum-CODE^@^ system was used. Each Spectrum-CODE bead was respectively pre-conjugated with a unique fluorophore combination selected from the following 7 fluorophores: quantum dot 547 nm (Qdot547), Qdot572, Qdot604, Qdot641, AlexaFluor488, AlexaFluor552, or AlexaFluor638, providing a digitized spectrum that can be used to decode the RNA transcript by fluorescence imaging. In this framework, “1” was used to denote the inclusion and “0” to denote the exclusion of a fluorophore on a Spectrum-CODE bead, providing a 7-digit spectral code designated to one RNA transcript. In this way, the 7-channel fluorescent system is expanded to encode more than 100 transcripts (maximum capacity, 2^7^) in one imaging cycle (**Supplementary Figure S4**). The Spectrum-CODE beads were dispersed in 1% BSA solution containing 0.1% triton-X 100 solution, and stored at 4 □ for further usage.

### Fluorescence microscopy

The fluorescence images were captured with confocal laser scanning microscope (Leica SP8, 63× oil immersion objective, NA 1.4, Leica Microsystems). Briefly, after sampling, the biochips were removed from forceps and immersed in RNALater (Beyotime). Before imaging-based analysis, the DNA-encoded reporter sequences were incubated with the biochips for 30 minutes followed by extensive PBS rinse Fluorescence images were acquired using a confocal laser scanning microscope. All the imaging parameters, including the laser power, gain, and scanning speed were fixed across the microscopy process. Scanning was performed with a 1 μm step size along the Z-axis spanning the whole layer. For data collection from a biochip, imaging-based decoding was performed according to the spectrum encoding (**Supplementary Table S1**). To acquire the whole view of a biochip, we used the navigator mode in Leica Microsystems to capture over 200 tiles for single biochip. The resulting large-scale tiles were merged using Leica imaging software (Leica Application Suite X, LAS X).

### Generation of RNA expression matrix

To transform the fluorescence image to matrix, ImageJ, python, and R were used. The identification of Spectrum-CODE beads was performed in Python. For analyzing the expression of an RNA transcript, the fluorescence image of a biochip undergoes processes involving binarization, mask generation, nanoprobe ROI selection, denoising and spectral vector composition by extracting the signal from all 7 florescence channels. The featured spectral vectors were decoded by using a machine learning model to derive the Spectrum-CODE associated with the nanoprobes, which were counted to indicate the molecular abundance for different transcripts. The machine learning model was pre-established by using the training dataset consisting of the actual fluorescence patterns collected from all possible Spectrum-CODEs. For each one, >200 samples were collected from multiple biochips to construct the training dataset.

Specifically, the fluorescence images were acquired as 3-dimensional stack (x, y, z), which were cropped to remove unnecessary edges in ImageJ, and then loaded in Python by using OpenCV package. For each nanoprobe ROI, the associated Spectrum-CODE puncta were identified by using *connectedcomponents* function in Python; recognized by *randomforest* to determine the corresponding SpectrumCODE, which was then counted. The counts of all 55 Spectrum-CODEs associated with every nanoprobe across a whole biochip were used to compose the RNA expression matrix for the ENDO-Genome tissue sampling site, which was later undergoes spatial transcriptomic analysis. In the analytical protocol, to explore the spatial transcriptomics association with CD, we defined quasi-single-cell spots (qSC-spots) by taking 4×4 binning of the nanoprobes.

### Analysis of CD Active score (related to Figure 5)

The data set contains 140,000 qSC-spots derived from >2000000 nanoprobes using 4×4 binning and was analyzed by unsupervised data clustering analyzed by Leiden algorithm to derive the qSC-spot clusters. qSC^(k)^ represents one of overlapping qSC-spot clusters, which were identified to have significant association with the “active” CD labeling or with the transcriptomic sampling at IL. The reference vector f^(k)^ is derived by averaging the expression data (***E***) associated all qSC spots in the cluster:

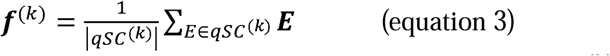

m is the number of overlapping clusters. The weighting coefficient W^(k)^ for cluster k is derived by calculating the portion of its qSC-spots out of the total number of all qSC-spots for the overlapping clusters, which is:

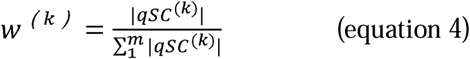

When applied to all the qSC-spot data, for a single qSC-spot expression vector E, the Euclidean distance d^(k)^ between E and each reference vector f^(k)^ was calculated:

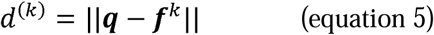

The W^(k)^ weighted distance D for a qSC-spot is calculated, the Active score (A-score) is derived by taking the reciprocal of the weighted distance:

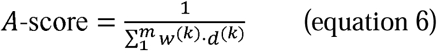

### Analysis of distance-dependent transcriptomic patterning for gene-pairs (related to Figure 6)

Least action distance (LAD) between anchor genes to target genes: For a specific ENDO-Genome biochip used for a patient, a selected gene a has ***m*** non-zero expression qSC-spots, which was defined as the anchor points. For each anchor point, its closest non-zero qSC-spot for a targeted paring gene, **b**, was determined as the *LAD_i_*:

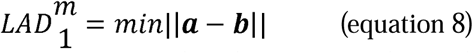

Accompanied expression probability (AEP) for gene **a** (anchor) and ***b*** was defined as the portion of short LAD (<20 μm, AEP_<20_) or long LAD (>80 μm, AEP_>80_) count out of all the **m** qSC-spots for in gene **a**:

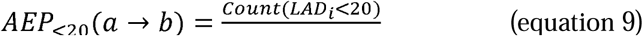

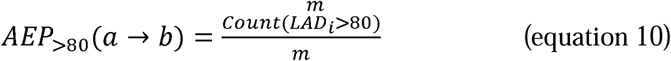

### Bioinformatics Analysis

Differential gene expression analysis (related to Figure 4) was conducted using the Seurat package. The *FindAllMarkers* function from Seurat was used to identify differentially expressed genes between clusters. This function uses a non-parametric Wilcoxon rank sum test to assess the differences in gene expression between groups. The analysis results were then filtered to identify the genes with a log-fold change greater than 1 and a p-value less than 0.05, determined as genes with substantial and statistically significant expression differences.

For analysis related to Figure 4 and 5, *Scanpy* in Python was used to analyze the mRNA expression data derived from over 2,000,000 nanoprobes (binned into 140,000 quasi - single - cell spots, namely qSC - spots, via 4×4 binning) obtained from 47 sampling studys across the 17 subjects. The UMAP (Uniform Manifold Approximation and Projection) algorithm was applied to visualize the mRNA expression data by embedding samples into a low dimensional space to show the sample clustering based on transcriptional expression. Unsupervised clustering analysis of the mRNA expression data from the 140,000 qSC - spots was also performed using *Scanpy*. Based on the multidimensional space formed by the 55 selected mRNA transcripts, the clustering algorithm in *Scanpy* was utilized to process the mRNA expression data, leading to the identification of 10 qSC-spot clusters (C1-C10). To visualize and compare these clustering patterns, the UMAP embedding method was used to reduce the dimensionality of the mRNA expression data, enabling a clearer observation and comparison of the distribution differences among different clusters in a two-dimensional space.

### Statistics and producibility

At least three independent biological replicates were used for all experiments. Statistical significance for comparing different experimental conditions was calculated by appropriate hypothesis test as indicated in the caption of each Figure. A p-value less than 0.05 was considered to be statistically significant, all the p-values are indicated in the respective Figures.

## Acknowledgement

This work was supported by National Natural Science Foundation of China (U20A20194), by General Research Fund (11215920, 11220024, 11218522, 11218523) from the Research Grants Council of Hong Kong SAR, and Shenzhen-Hong Kong-Macau Science and Technology Program (Category C, SGDX2020110309300502). Support from Innovation and Technology Commission of Hong Kong through the Centre for Cerebro-Cardiovascular Health Engineering and funds from City University of Hong Kong (7005084, 7005206, 7005642, 7020003, 7020077, 9680233, 9240060) are also acknowledged.

## Author Contribution

P.S. conceived the project and supervised the research. H.S, F.G. and P.S. designed experiments. H.S. and F.G. carried out the investigation and analyzed the data. Y.W. X.H., C.X., Y.M., J.Q., X.Z. and X.J. contributed to different aspects of the experimental works. Z.Z., X.Z., J.S. and J.K. carried out the clinical study. P.L. and Z.Y. contributed to the sensor design. B.G. and F.G. contributed to the data analysis. L.Y. M. and Y.H.Y. contributed to the clinical research design. H.S., F.G. and P.S. wrote the manuscript. J.K. L.Y. M. and Y.H.Y. contributed to the writing of the manuscript.

## Data availability

The data that support the findings of this study are available from the corresponding author on request.

