## Supplementary Materials for "In Vivo Spatial Transcriptomics for Bleeding-free Profiling Human Internal Organs"

**Supplementary Figures**

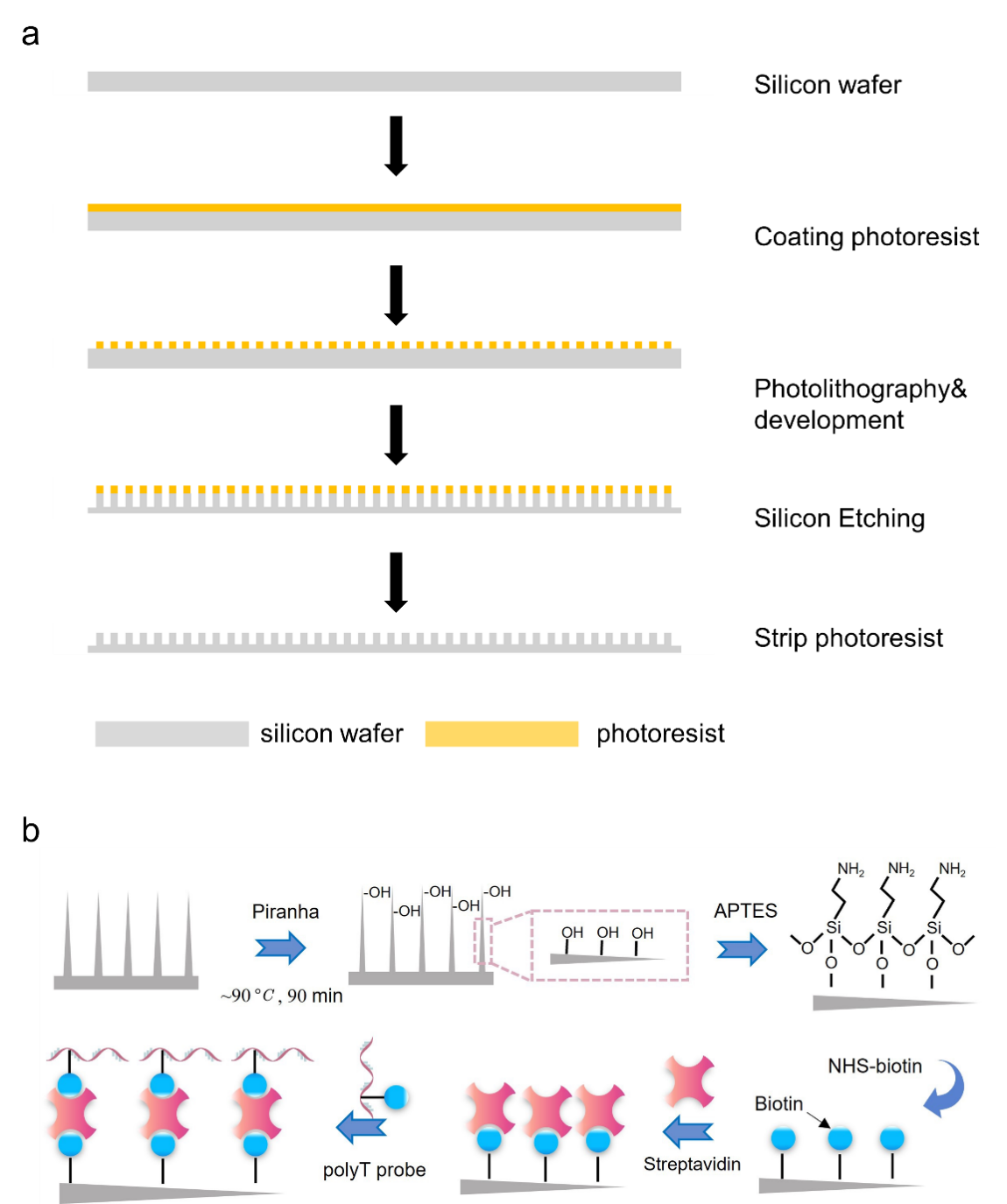

**Supplementary Figure S1. Fabrication and functionalization of the ENDO-Genome biochip. (a)** Fabrication of biochips with vertically aligned nanoprobes by deep reactive ion etching (DRIE) and regular photolithography. **(b)** Procedures of biochip functionalization of biochip. The biochip is first activated by piranha solution (H_2_SO_4_:H_2_O_2_=3:1). Then, -NH_2_ is introduced by 3-aminopropyltriethoxysilane (APTES) treatment. Biotin, streptavidin, and polyT DNAs are sequentially linked to the nanoprobes with proper binding chemistry.

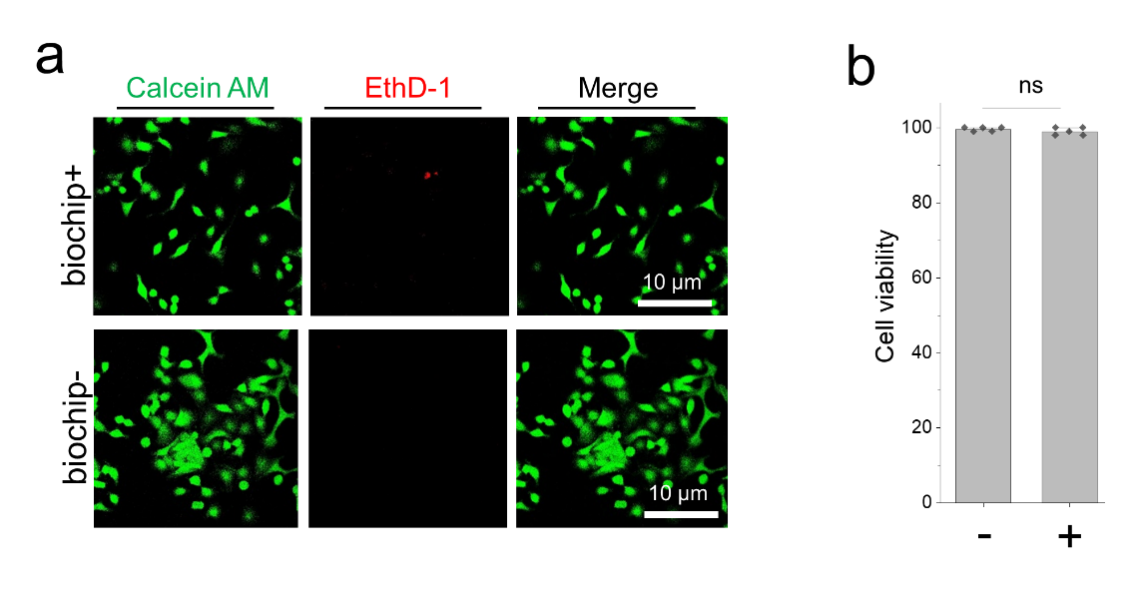

**Supplementary Figure S2. In vitro biocompatibility characterization of the ENDO-Genome biochip. (a)** Fluorescence images showing live (green, Calcein AM) and dead (red, EthD-1) cells (A549) after cellular interfacing with a ENDO-Genome biochip. The comparison groups are cells treat with (biochip+) or without (biochip-) a biochip. Scale bars, 10 μm. **(b)** Quantification of cell viability after biochip treatment. Data are presented as mean$\pm$SD, analyzed by two-tail student t-test.

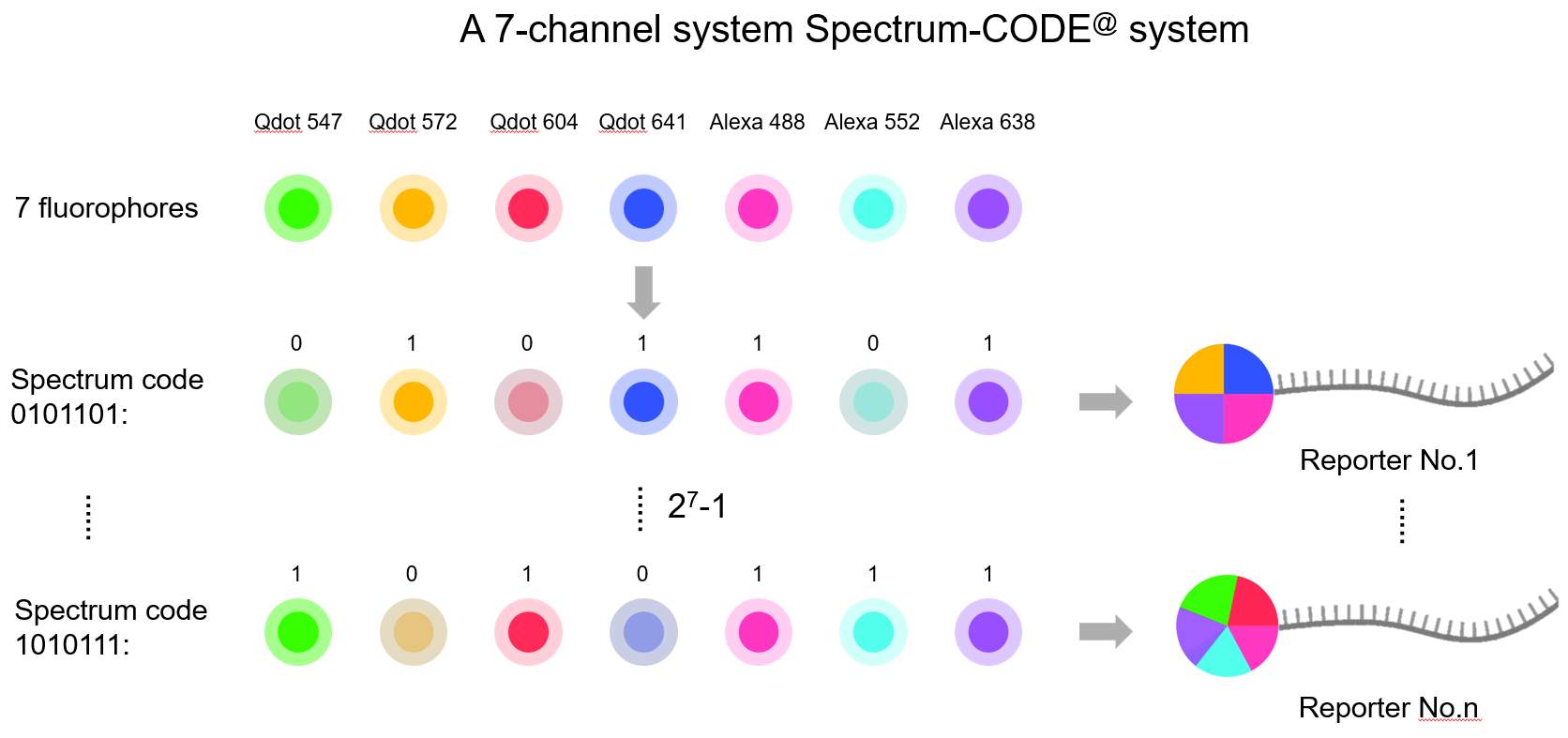

**Supplementary Figure S3. A 7-channel Spectrum-CODE^@^ system.** Each Spectrum-CODE bead was respectively pre-conjugated with a unique fluorophore combination, selected from the following 7 fluorophores: quantum dot 547 nm (Qdot547), Qdot572, Qdot604, Qdot641, AlexaFlor488, AlexaFlor552 or AlexaFlor638, providing a digitized spectrum that can be used to decode the RNA transcript by fluorescence imaging.

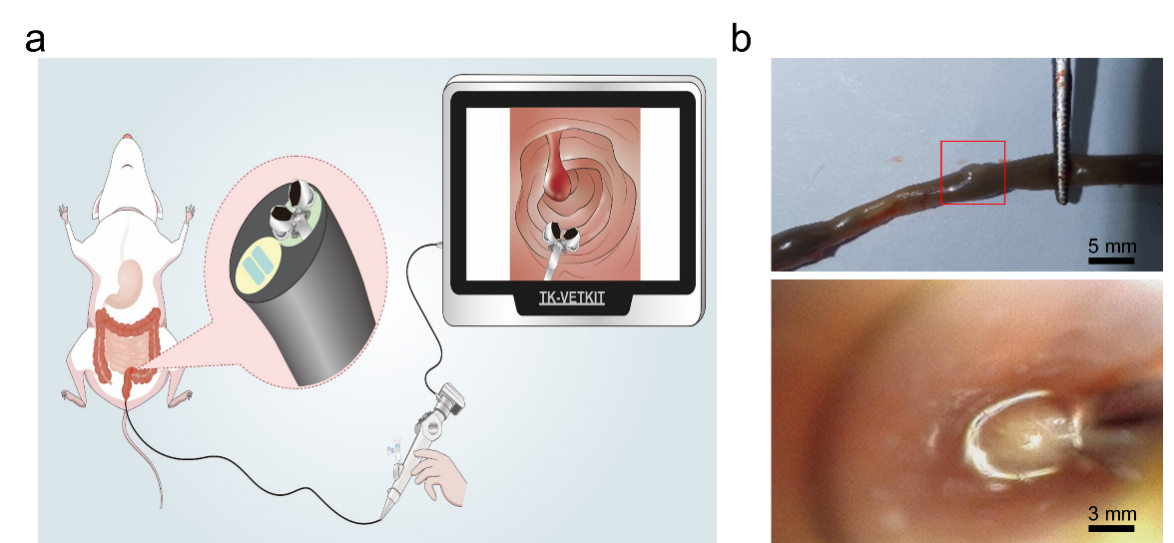

**Supplementary Figure S4.** **(a)** Schematic of ENDO-Genome operation in a mouse model. **(b)** Top: optical image of polyps area (red rectangle) of extracted mouse colon, scale bar, 5 mm; bottom: in body colonoscopy image of polyps area in the mouse colon, scale bar, 3 mm.

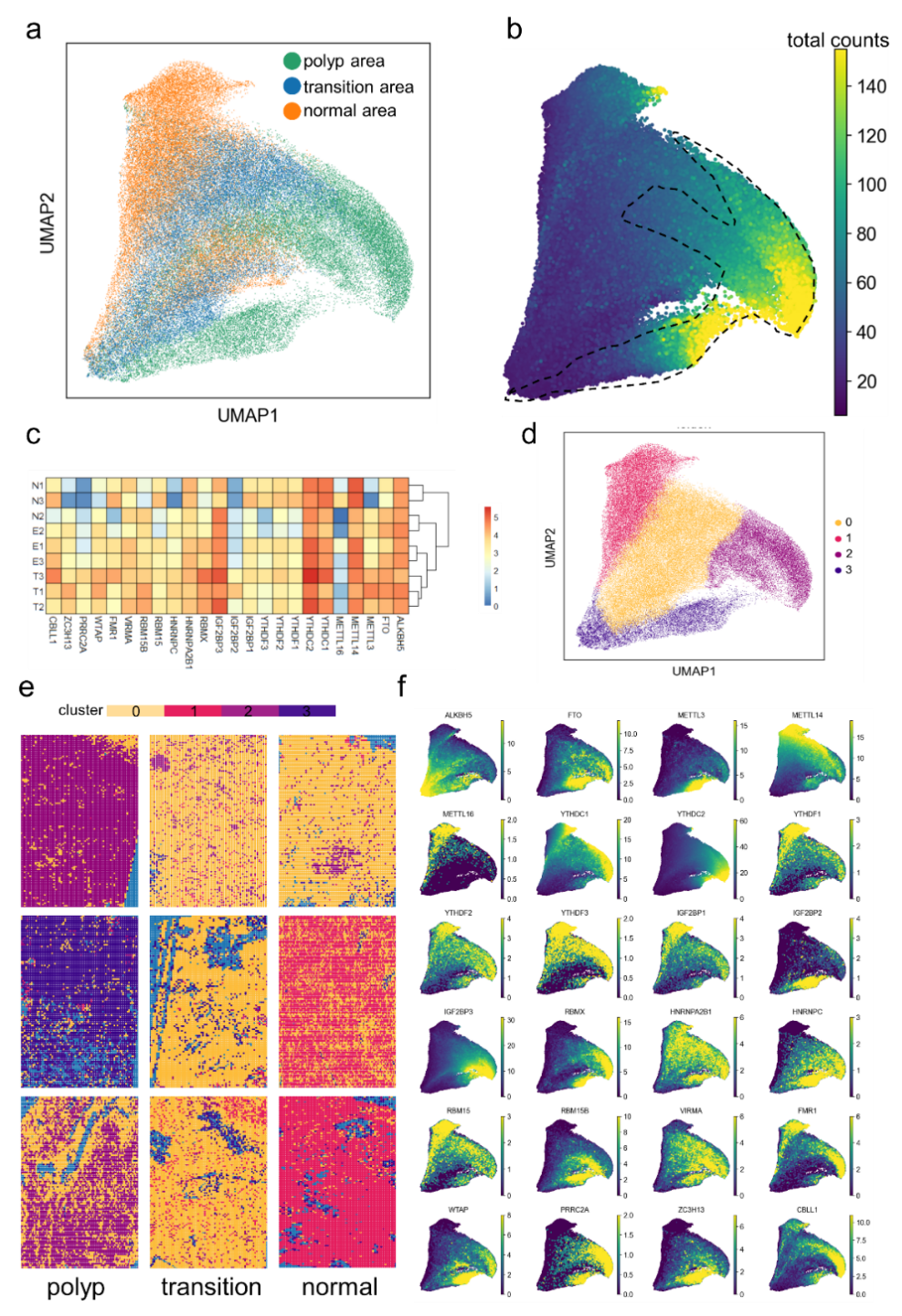

**Supplementary Figure S5.** ***In vivo* spatial transcriptomics profiling in mouse cancer model using ENDO-Genome. (a)** UMAP embedding of over 80,000 qSC-spots (4x4 binning from over 1000000 nanoprobes) marked by different sampling area (green for polyp area, blue for transition area, and orange for normal area). Each point represents the two-dimensional projection of an individual qSC-spot associated with expression of 24 mRNAs, illustrating the intrinsic coherence based on gene expressions. **(b)** UMAP that indicates the gene expression counts among the two-dimensional cluster space, with lighter colors indicating more count at this space. The polyp-like cluster marked in dotted line presents higher gene expression than other clusters. **(c)** Heatmap of average expression for the RNA methylation gene set across 9 samples. Rows represent individual samples, while columns represent genes. **(d)** UMAP that indicates the unsupervised three clusters. **(e)** The three unsupervised clusters are projected on biochip as spatial scatters. Cluster 2 and 3 are enriched in polyp samples, cluster 0 is enriched in transition samples, and cluster 1 is enriched in normal samples. **(f)** UMAP indicated the enrichment of each mRNA among the two-dimensional cluster space, with lighter colors indicating higher count in this cluster.

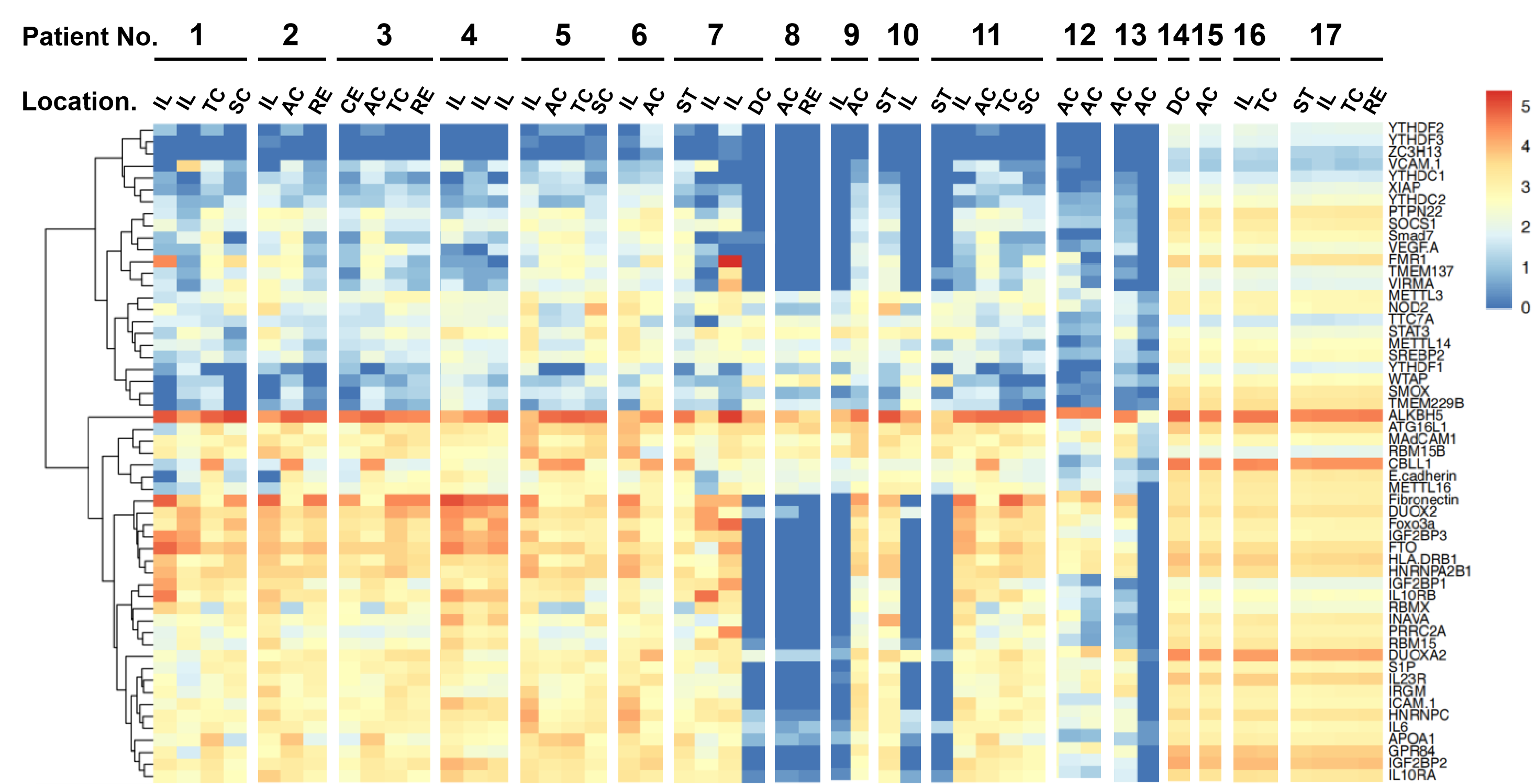

**Supplementary Figure S6.** Heatmaps of average expression for the CD gene set across 17 subjects and 8 GI location. Rows represent genes, while columns represent individual samples.

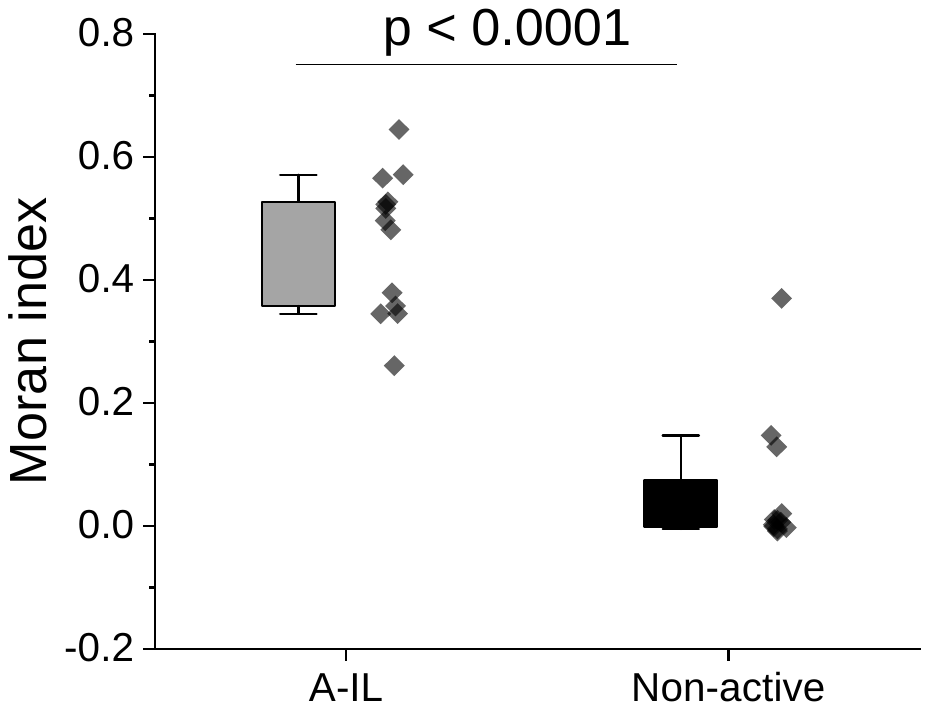

**Supplementary Figure S7.** Boxplot with scattered data points showing the comparison of the Moran index comparison for samples of active ileum sampling versus all non-active sampling (remission and healthy subjects). The black dots indicate an overlay of individual data points for each sample, the box indicates the first quartile, median, third quartile of the data, the whisker limit is 1.5 times the interquartile range, analyzed by Kruskal-Wallis test.

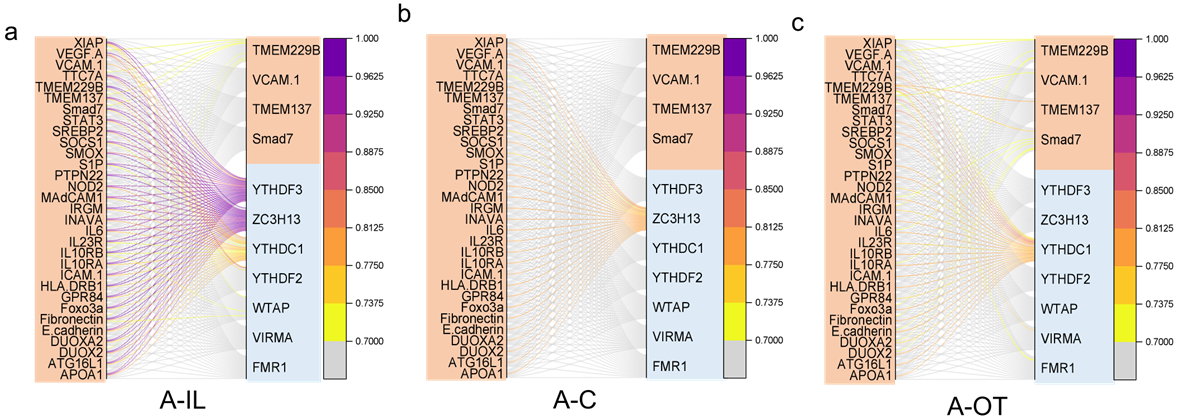

**Supplementary Figure S8.** Analysis of distance-dependent transcriptomic patterning for gene-pairs at multi-cell scale (>80 μm) in active CD patients. Comparison is made among A-IL (**a**), A-C (**b**) and A-OT (**c**) categories. Mapping from the anchor genes (left column), the spatial distance of the closest expression of the pair gene (right column) was prioritized in the inflammation (brown) or methylation (blue) regulator groups. The associative probability was indexed by a colored line connecting the anchor and the pair genes with a threshold set at 70%.

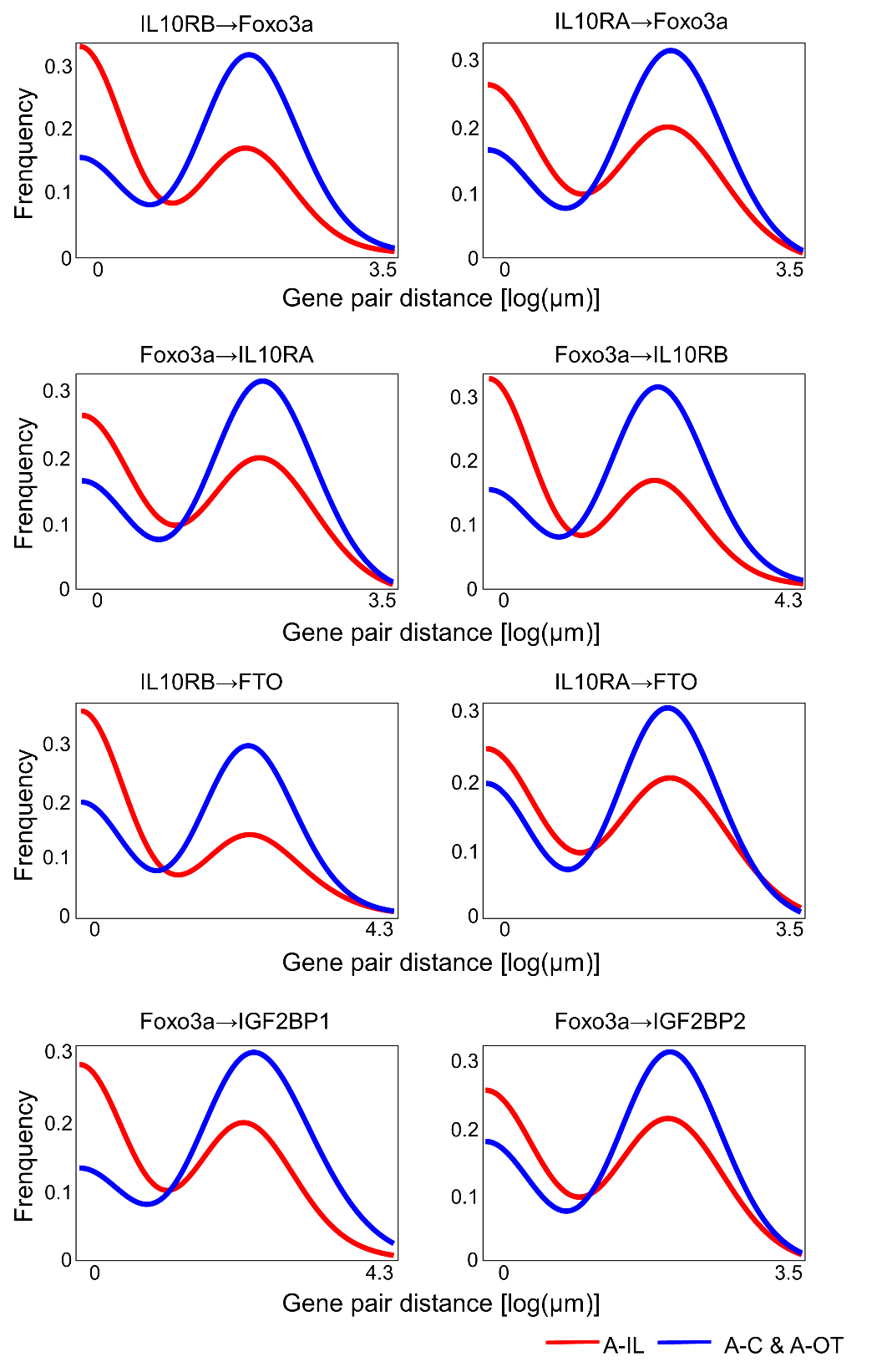

**Supplementary Figure S9**. Histogram of the association distance for representative gene pairs, comparing inflammation-responsive participation of endothelial dysregulation or RNA methylation related genes in the ileum (red curve) of active CD patients.

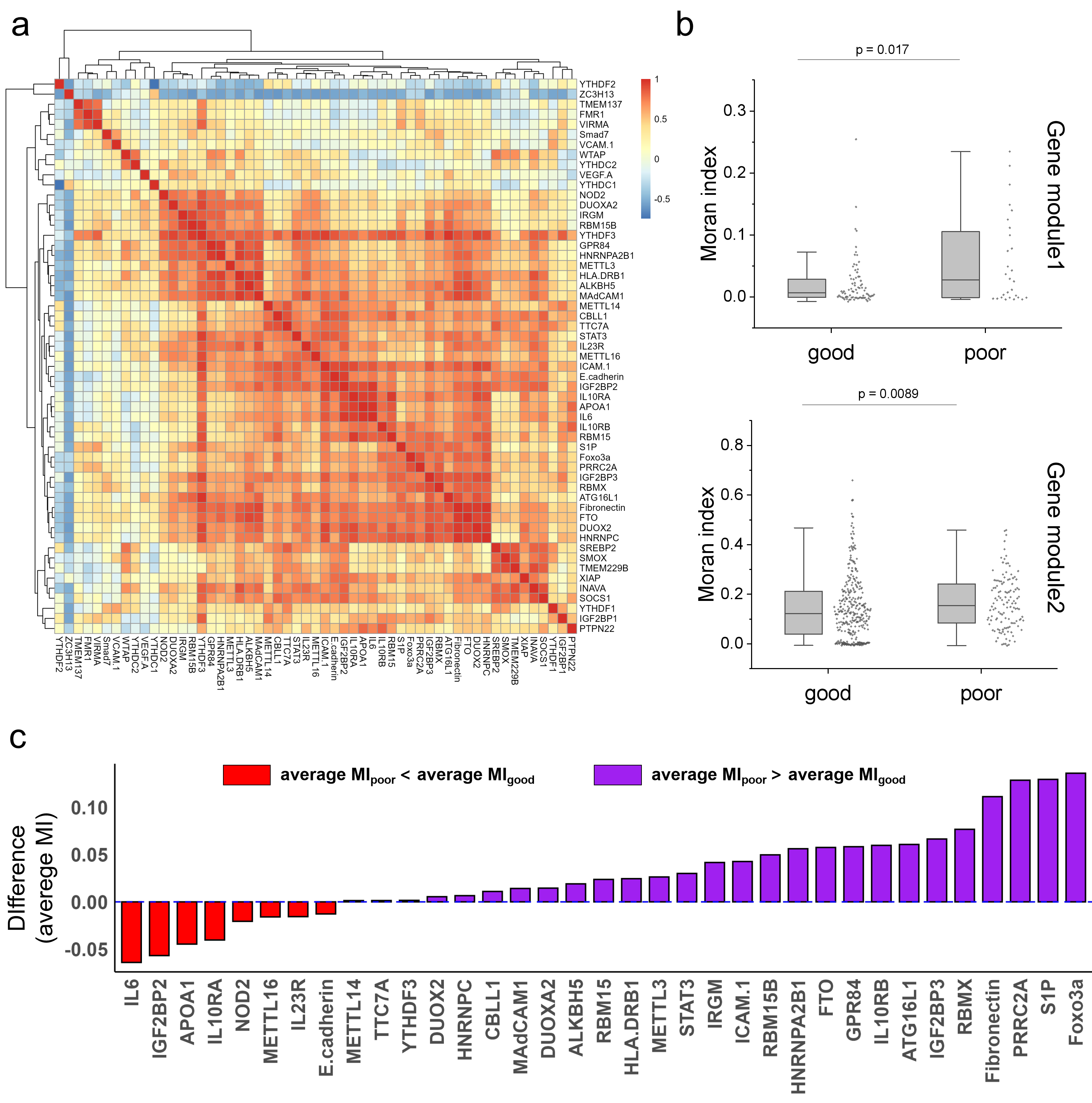

**Supplementary Figure S10. The association between Moran index (MI) of genes and prognosis status among CD patients.** (**a**) Correlation of genes determined by their MI over 13 CD patients, with red indicating positive correlation and blue indicating negative correlation. The correlation coefficients range from -1 to 1, and the color legend clearly shows the correspondence between correlation coefficients and colors. The gene clustering dendrogram reveals distinct gene modules. (**b**) Boxplots showing the individual gene MI from different gene modules, pooled from 13 Crohn's disease patients with different prognosis (good prognosis and poor prognosis). Each box indicates the first quartile, median, third quartile of the data, the whisker limit is 1.5 times the interquartile range, analyzed by Kruskal-Wallis test. (**c**) Barplots showing the difference of the average gene MI between poor prognosis group and good prognosis group.
**Supplementary Tables**

**Supplementary Table S1.** List of gene encoding and associated reporter sequences.

| **Gene target** | **Encoded Reporter Sequence (5' to 3')** | **Spectrum code** |
| --- | --- | --- |
| YTHDF1 | AATGACTTTGAGCCCTACCTTTCTGGACAGTCCAAT | 0001100 |
| YTHDF2 | CCAGCAGCCTCTTGGAGCAGAGACCAAAAGGTCAAG | 0100001 |
| YTHDF3 | GACAGGCCGGATTTGGCAATGATACTTTGAGTAAGG | 0011010 |
| YTHDC1 | GATTTCTAAGCCACTAAGCTCATCTGTAAGCAATAA | 0000110 |
| YTHDC2 | CTGGAGCGCTTCCGCTACGGTGACCAGAGAGAAATG | 0000100 |
| HNRNPC | AAGGCTTTGCCTTTGTCCAGTATGTTAATGAAAGAAATGCC | 0010110 |
| RBMX | GGTGGGCTTAATACAGAGACGAATGAGAAAGCCCTT | 0101010 |
| HNRNPA2B1 | ACCACAGAAGAAAGTTTGAGAAACTACTATGAGCAATGGGG | 0100000 |
| IGF2BP1 | ATGAACAAGCTTTACATCGGCAACCTCAACGAGAGT | 0010101 |
| IGF2BP2 | ATGATGAACAAGCTGTACATTGGGAACCTGAGCCCC | 0100010 |
| IGF2BP3 | ACTGAATGGATTCCAGTTAGAGAACTTCACCTTGAAGGTTG | 0010111 |
| FMR1 | CGGGGCTCCAATGGCGCTTTCTACAAGGCATTTGTA | 0100101 |
| PRRC2A | GGGCCGACTGCCAAGGGGAAGGATGGAAAGAAGTAT | 0100011 |
| METTL3 | ATGTCGGACACGTGGAGCTCTATCCAGGCCCATAAG | 1001100 |
| METTL14 | AATAGCAAAGATGAACAGAGGGAGATTGCTGAAACCAGA | 0101111 |
| METTL16 | TCAATGCATGCAAGAAATAGATACAAGGACAAACCACCTGA | 0101110 |
| WTAP | TAGTGAGGAGCTTAAAAGCAGTCAGGATGAACTGAATGACT | 1001001 |
| VIRMA | TTAGATACTTTTAAACATCCGAGCGCTGAGCAAAGTTCTCA | 0000111 |
| ZC3H13 | AAAATTAGGAGAAAGGTCACAGTGGAAAACACCAAGACCA | 0101011 |
| CBLL1 | GACAATGAGTTACAAGGCACTAATAGTTCTGGATCCTTGGG | 0101100 |
| RBM15 | GACGGCGTGGAGTACAAGACCCTGAAGATCAGCGAG | 0011101 |
| RBM15B | GAGGACCTGATGCCGGAGGATGATCAGAGAGCCACT | 1100111 |
| FTO | CGCGGAGGAACGAGAGCGGGAAGCTAAGAAACTGAG | 0011001 |
| ALKBH5 | CTACACCGACCTGCGGGAGAAGCTCAAGTCCATGAC | 0101000 |
| MAdCAM1 | CCACGCAGGGAGAAGTGATCCCAACAGGCTCGTCCAAA | 1000001 |
| XIAP | TTATGAAGCACGGATCTTTACTTTTGGGACATGGATAT | 0000101 |
| GPR84 | CCAAGCTCCGTACCCGATTCAACCTGCTCATAGCCAACC | 0110000 |
| Smad7 | CGGATCTGAAGGCGCTCACGCACTCGGTGCTCAAGAAA | 0011000 |
| Foxo3a | TCTACAGCAGCTCAGCCAGCCTGTCACCTTCAGTAAGCAA | 1100001 |
| TMEM229B | CTGTATGCCATCCACGGCTACTTCTGCGAGGTGATGTTCA | 0001000 |
| IL10RA | CCTCCACTGGACACCCATCCCAAATCAGTCTGAAAGTACC | 0010011 |
| IL10RB | CTTGACGGAATGTGATTTCTCAAGTCTTTCCAAGTATGGT | 0010100 |
| IL23R | TTCCTGTGAAATGAGATACAAGGCTACAACAAACCAAACT | 0000001 |
| IL6 | GTTGCCTTCTCCCTGGGGCTGCTCCTGGTGTTGCCTGCTG | 0011100 |
| STAT3 | ACGTCATTAGCAGAATCTCAACTTCAGACCCGTCAACAAA | 0010000 |
| SOCS1 | CGACACGCACTTCCGCACATTCCGTTCGCACGCCGATTAC | 0001011 |
| APOA1 | GACACGCACTTCCGCACATTCCGTTCGCACGCCGATTACC | 1010001 |
| DUOXA2 | CATGCCGCAGGCTTCAGCGTTCCACTGCTCATCGTTATTC | 0011110 |
| DUOX2 | AGAAGGAAGCAGCCAAAGATGGAGTGCCAGCGATGGAGTG | 0011111 |
| TMEM137 | GAGGAGCTGGCAGCCCTCTTTGCGCCCTACGGCACGGTCA | 0101001 |
| TTC7A | AGAAGACCACAAATAACAGCACGTCGAGGCATCTGAAAGG | 0100111 |
| SMOX | GCATCAGCCTCTATTCCAAGAATGGCGTGGCCTGCTACCT | 0000011 |
| ICAM-1 | GGAACAACCGGAAGGTGTATGAACTGAGCAATGTGCAAGA | 0110001 |
| VCAM-1 | TATCTTGCTCAGATTGGTGACTCCGTCTCATTGACTTGCA | 0000010 |
| Fibronectin | GACTGCTCAACAGACAACCAAACTGGATGCTCCCACTAAC | 0101101 |
| E-cadherin | CAGGTGGGATGAGGATGAGCAGTGGTCGGTGGTGGTGGAG | 0100110 |
| VEGF-A | TCCAGGAGTACCCTGATGAGATCGAGTACATCTTCAAGCC | 0001001 |
| SREBP2 | AAGTCCTTCAGCCTCAAGTCCAAAGCCTGGTGACATCCTC | 0001110 |
| S1P | GTATGAACTGAGCAATGTGCAAGAAGATAGCCAACCAATGTG | 0100100 |
| ATG16L1 | CAGATAGATAGTCCACTGAATGGGAAGGTGACGAATGAGG | 0010010 |
| HLA-DRB1 | CAGCACGTACCTCTATAACCGCCAGCGGAAGATCAAGAAA | 0001111 |
| INAVA | AGGAGAACCTCAGCAGGCAGGCTCGGCGGCAGCGGAAGCA | 0011011 |
| IRGM | GTCTGCCACCACAACCCTGGAGAACTACCTGATGGAAATG | 0010001 |
| NOD2 | ACCTCAATGACGATGCGGACACTGTGCTGGTGGTGGGTGA | 0001010 |
| PTPN22 | GCTGGCTGTGGAAGGACTGGTGTTATTTGTGCTATTGATT | 0001101 |

**Supplementary Table S2.** Annotations of all mRNA transcripts profiled in this study.

| **mRNA** | **Function**  **group** | **Functional relevance to CD ^#^** |
| --- | --- | --- |
| YTHDF1 | m6A RNA methylation reader | stem cell homeostasis, intestinal barrier integrity ^1^ |
| YTHDF2 |  | intestinal epithelial cell proliferation and apoptosis ^2,3^ |
| YTHDF3 |  | mRNA translation and degradation ^1,3^ |
| YTHDC1 |  | inflammatory responses, epithelial barrier integrity ^4^ |
| YTHDC2 |  | RNA metabolism ^5,6^ |
| HNRNPC |  | cell chemotaxis and migration ^6,7^ |
| RBMX |  | RNA splicing and metabolic regulation ^5,8^ |
| HNRNPA2B1 |  | macrophage polarization, mRNA stability of pro-inflammatory cytokines ^8,9^ |
| IGF2BP1 |  | maintains intestinal barrier function by stabilizing occluding mRNA ^10^ |
| IGF2BP2 |  | macrophage conversion, ferroptosis ^6,8,11^ |
| IGF2BP3 |  | RNA stability and translation ^11,12^ |
| FMR1 |  | m6A regulation ^6,8,13^ |
| PRRC2A |  | epithelial cell regeneration and repair ^6,14^ |
| METTL3 | m6A RNA methylation writer | inflammatory response, signaling transduction ^6,13,15^ |
| METTL14 |  | stem cell apoptosis, mucosal barrier function ^6,13^ |
| METTL16 |  | m6A modification of U6 snRNA and mRNAs ^6,8^ |
| WTAP |  | key component of the m6A writer complex ^6,13,16^ |
| VIRMA |  | m6A modification, inflammation-related gene expression ^1,6,8^ |
| ZC3H13 |  | intestinal epithelial cell regeneration and repair ^1,6,8^ |
| CBLL1 |  | immune cell signaling, cytokine production ^3,6,8^ |
| RBM15 |  | inflammation regulation, immune response ^1,6,8^ |
| RBM15B |  | intestinal homeostasis, inflammatory responses ^1,6,8^ |
| FTO | m6A RNA methylation eraser | immune response, signaling transduction and autophagy ^1,8,13^ |
| ALKBH5 |  | cytokines, cell proliferation, apoptosis, and barrier integrity ^1,2,6^ |
| MAdCAM1 | inflammation regulation | lymphocyte recruitment to the intestinal mucosa ^17,18^ |
| XIAP |  | cytokines, signaling transduction, apoptosis ^19-21^ |
| GPR84 |  | pro-inflammatory responses in immune cells ^22-24^ |
| Smad7 |  | immune response, cytokines, signaling transduction ^25,26^ |
| Foxo3a |  | inflammatory responses in monocytes, cytokines ^27,28^ |
| TMEM229B |  | cellular signaling, membrane transport, or cell-cell interactions ^29^ |
| IL10RA |  | immune response, signal transduction, cytokines ^30-32^ |
| IL10RB |  | immune response, signal transduction, cytokines ^32,33^ |
| IL23R |  | JAK-STAT signaling, pro-inflammatory cytokine production ^34^ |
| IL6 |  | pro-inflammatory, T cell activation ^35^ |
| STAT3 |  | adaptive immunity, cytokines ^36^ |
| SOCS1 |  | immune tolerance in the intestine ^37^ |
| APOA1 |  | anti-immune response ^38^ |
| DUOXA2 |  | thyroid hormone synthesis, immune regulation ^39^ |
| DUOX2 |  | reactive oxygen species in the intestine ^39^ |
| TMEM137 |  | cytokine production, innate immune response ^40^ |
| TTC7A | cell proliferation | cell differentiation, immune response ^41^ |
| SMOX |  | microbiome homeostasis ^42^ |
| ICAM-1 |  | cell proliferation, apoptosis, signal transduction ^43^ |
| Fibronectin |  | adhesion, migration, signaling transduction ^44^ |
| SREBP2 |  | signaling transduction ^45^ |
| VCAM-1 |  | cytokines, signal transduction, cell adhesion ^46^ |
| E-cadherin |  | integrity of the intestinal barrier ^47^ |
| VEGF-A |  | cell adhesion, cytokines, immune response ^48^ |
| S1P |  | cytokines, signaling transduction ^49^ |
| ATG16L1 |  | anti-bacterial autophagic response, cytokines ^50^ |
| HLA-DRB1 |  | adaptive immunity ^51^ |
| INAVA |  | cytokine, signaling transduction ^52^ |
| IRGM |  | anti-bacterial autophagic response ^53^ |
| NOD2 |  | Immune regulation, susceptibility to inflammation ^20,31^ |
| PTPN22 |  | cytokine production, immune cell activation ^31^ |

**Supplementary Table S3.** Information summary for subjects involved in human clinical study.

| **Subject**  **No.** | **Sample**  **No.** | **Age** | **Sampling location** | **CD Status** | **SES-CD ^#^** |
| --- | --- | --- | --- | --- | --- |
| 1 | 1 | 45 | stomach | active | 4 |
| 1 | 2 | 45 | ileum | active | 4 |
| 2 | 3 | 34 | ileum | active | 3 |
| 2 | 4 | 34 | ascending colon | active | 4 |
| 3 | 5 | 44 | ascending colon | active | 4 |
| 3 | 6 | 44 | rectum | active | 4 |
| 4 | 7 | 32 | sigmoid colon | remission | 2 |
| 5 | 8 | 28 | ascending colon | remission | 1 |
| 6 | 9 | 32 | stomach | active | 3 |
| 6 | 10 | 32 | ileum | active | 4 |
| 6 | 11 | 32 | ileum | active | 4 |
| 6 | 12 | 32 | descending colon | active | 3 |
| 7 | 13 | 41 | ileum | active | 4 |
| 7 | 14 | 41 | ileum | active | 4 |
| 7 | 15 | 41 | transverse colon | active | 3 |
| 7 | 16 | 41 | sigmoid colon | active | 4 |
| 8 | 17 | 50 | ileum | active | 3 |
| 8 | 18 | 50 | ascending colon | active | 4 |
| 8 | 19 | 50 | rectum | active | 4 |
| 9 | 20 | 23 | cecum | active | 3 |
| 9 | 21 | 23 | ascending colon | active | 3 |
| 9 | 22 | 23 | transverse colon | active | 4 |
| 9 | 23 | 23 | rectum | active | 4 |
| 10 | 24 | 23 | stomach | active | 3 |
| 10 | 25 | 23 | ileum | active | 4 |
| 10 | 26 | 23 | ascending colon | active | 4 |
| 10 | 27 | 23 | transverse colon | active | 4 |
| 10 | 28 | 23 | sigmoid colon | active | 3 |
| 11 | 29 | 27 | ileum | active | 4 |
| 11 | 30 | 27 | ileum | active | 4 |
| 11 | 31 | 27 | ileum | active | 4 |
| 12 | 32 | 33 | ascending colon | healthy | 0 |
| 12 | 33 | 33 | ascending colon | healthy | 0 |
| 13 | 34 | 43 | ascending colon | healthy | 0 |
| 13 | 35 | 43 | ascending colon | healthy | 0 |
| 14 | 36 | 26 | ileum | remission | 2 |
| 14 | 37 | 26 | transverse colon | remission | 1 |
| 15 | 38 | 52 | stomach | remission | 1 |
| 15 | 39 | 52 | ileum | remission | 2 |
| 15 | 40 | 52 | transverse colon | remission | 1 |
| 15 | 41 | 52 | rectum | remission | 1 |
| 16 | 42 | 20 | ileum | active | 3 |
| 16 | 43 | 20 | ascending colon | active | 4 |
| 16 | 44 | 20 | transverse colon | active | 4 |
| 16 | 45 | 20 | sigmoid colon | active | 4 |
| 17 | 46 | 23 | ileum | active | 3 |
| 17 | 47 | 23 | ascending colon | active | 3 |

^#^As part of clinical care, patients were assessed using the Simple Endoscopic Score for Crohn's Disease (SES-CD), evaluated across all segments. Subjects with an SES-CD $\geq$ 3 were classified as active, while those with an SES-CD $\leq$ 2 were considered in remission. Healthy subjects exhibited an SES-CD of 0.

**Supplementary Table S4.** Conditional gene pairing probability between CD inflammation genes and selected target genes over a short-distance scale (<20 µm).

| **Target gene** | **Function group** | **A-IL#** | **A-C#** | **A-OT#** |
| --- | --- | --- | --- | --- |
| ALKBH5 | RNA methylation | 0.97 | 1.00 | 1.00 |
| IGF2BP3 |  | 0.95 | 0.05 | 0.81 |
| FTO |  | 0.90 | 0.96 | 0.95 |
| HNRNPA2B1 |  | 0.87 | 0.94 | 0.92 |
| HNRNPC |  | 0.84 | 0.11 | 0.37 |
| IGF2BP2 |  | 0.83 | 0.80 | 0.78 |
| RBMX |  | 0.80 | 0.00 | 0.00 |
| IGF2BP1 |  | 0.72 | 0.00 | 0.00 |
| RBM15B |  | 0.36 | 0.05 | 0.00 |
| RBM15 |  | 0.15 | 0.00 | 0.00 |
| CBLL1 |  | 0.00 | 0.69 | 0.00 |
| METTL3 |  | 0.00 | 0.00 | 0.02 |
| Fibronectin | CD-related  Inflammation | 0.95 | 0.60 | 0.93 |
| DUOX2 |  | 0.91 | 0.89 | 0.53 |
| HLA.DRB1 |  | 0.87 | 0.75 | 0.90 |
| Foxo3a |  | 0.86 | 0.76 | 0.84 |
| ICAM.1 |  | 0.78 | 0.00 | 0.05 |
| IL10RB |  | 0.74 | 0.07 | 0.19 |
| IL10RA |  | 0.73 | 0.49 | 0.10 |
| MAdCAM1 |  | 0.66 | 0.00 | 0.48 |
| IL6 |  | 0.54 | 0.00 | 0.00 |
| ATG16L1 |  | 0.36 | 0.82 | 0.36 |
| STAT3 |  | 0.05 | 0.00 | 0.00 |
| INAVA |  | 0.05 | 0.00 | 0.00 |
| APOA1 |  | 0.05 | 0.43 | 0.00 |
| IL23R |  | 0.05 | 0.00 | 0.76 |
| DUOXA2 |  | 0.02 | 0.33 | 0.40 |
| GPR84 |  | 0.00 | 0.82 | 0.30 |
| IRGM |  | 0.00 | 0.00 | 0.17 |
| S1P |  | 0.00 | 0.00 | 0.40 |

\# A-IL, A-C, and A-OT are grouped by sampling locations at ileum, colon, or other locations (stomach and rectum) in active Crohn’s disease (CD) patients.

**Supplementary Table S5**. Conditional gene pairing probability between CD inflammation genes and selected target genes over a large-distance scale (>80 µm).

| **Target gene** | **Function group** | **A-IL^#^** | **A-C^#^** | **A-OT^#^** |
| --- | --- | --- | --- | --- |
| YTHDF3 | RNA methylation | 0.95 | 0.00 | 0.00 |
| ZC3H13 |  | 0.90 | 0.78 | 0.00 |
| YTHDC1 |  | 0.67 | 0.00 | 0.76 |
| YTHDF2 |  | 0.08 | 0.00 | 0.00 |
| WTAP |  | 0.07 | 0.00 | 0.00 |
| VIRMA |  | 0.00 | 0.00 | 0.02 |
| FMR1 |  | 0.00 | 0.00 | 0.05 |
| TMEM229B | CD related Inflammation | 0.16 | 0.00 | 0.12 |
| VCAM.1 |  | 0.00 | 0.00 | 0.02 |
| TMEM137 |  | 0.00 | 0.00 | 0.03 |
| Smad7 |  | 0.00 | 0.00 | 0.16 |

\# A-IL, A-C, and A-OT are grouped by sampling locations at ileum, colon, or other locations (stomach and rectum) in active Crohn’s disease (CD) patients.

**Supplementary Movies**

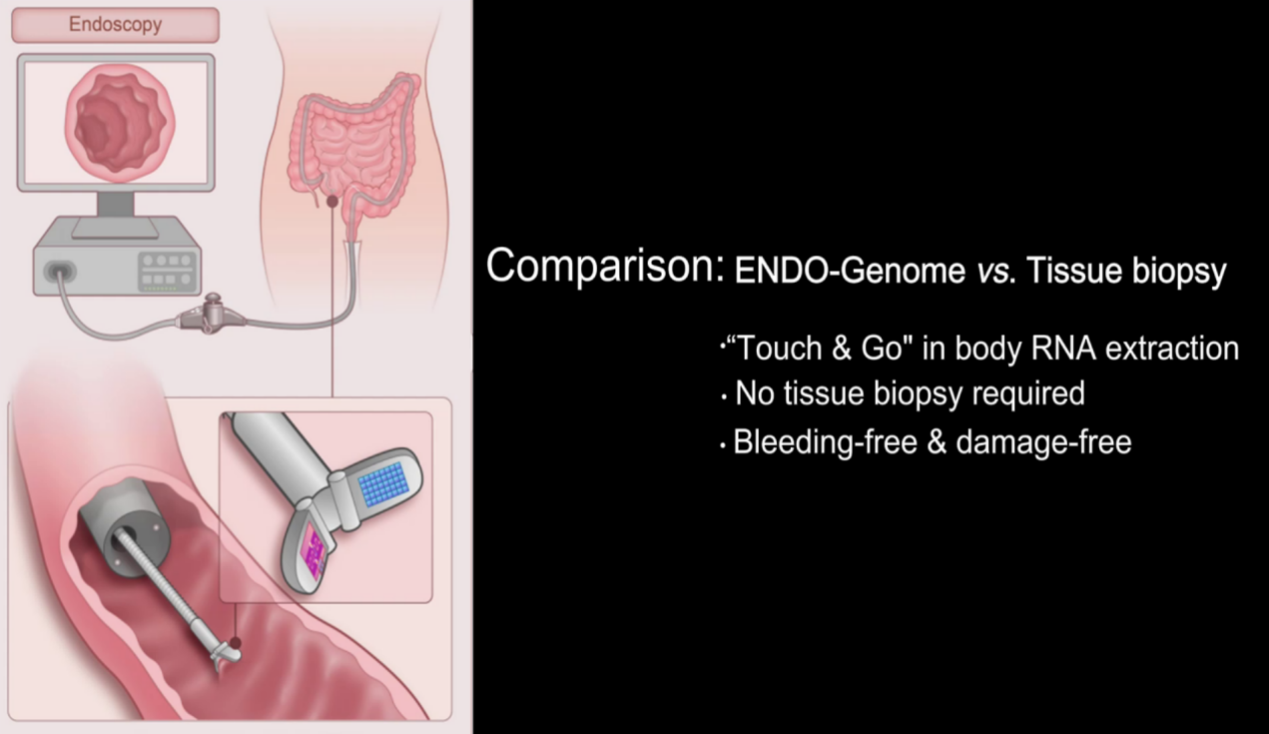

**Supplementary Movie S1.**	Bleeding comparison: ENDO-Genome VS. Tissue biopsy.

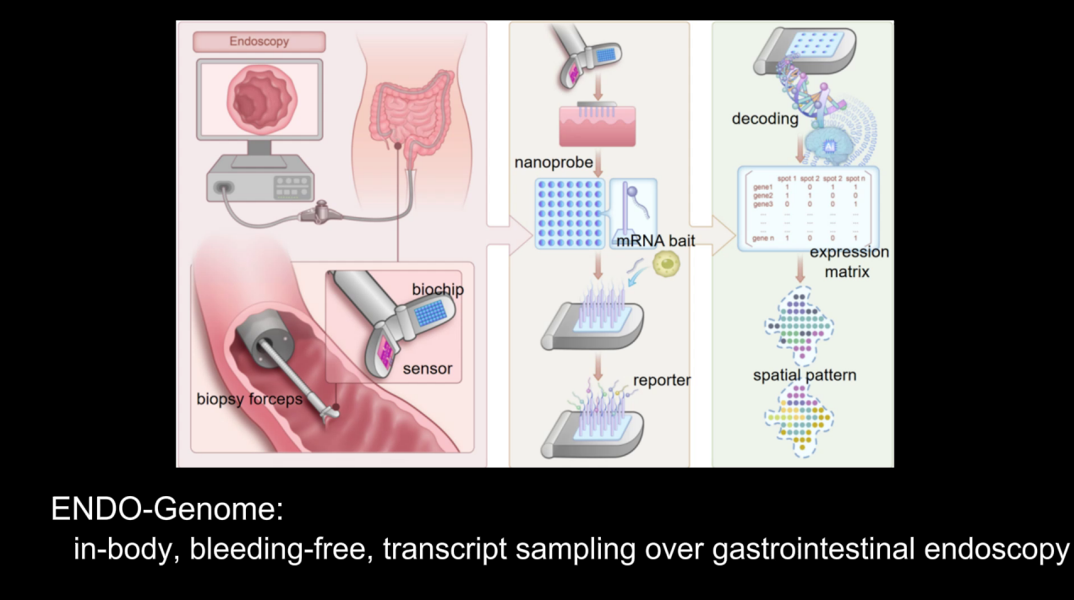

**Supplementary Movie S2.** ENDO-Genome: in-body, bleeding-free, transcript sampling over gastrointestinal endoscopy.
